# Myasthenia Gravis Activities of Daily Living Deep Phenotyping and Longitudinal

**DOI:** 10.64898/2026.10.26.26366495

**Authors:** Marc Garbey, Quentin Lesport, MGNet Investigators, Olivia Haddad, Charles Carpenter, Henry J. Kaminski, MGNet Investigators

**Affiliations:** Department of Neurology & Rehabilitation Medicine, George Washington University, Washington DC USA; Laboratoire des Sciences de l’Ingénieur pour l’Environnement (LaSIE) UMR-CNRS 7356 Université of La Rochelle, France; Care Constitution Corp, Houston, TX, USA

**Keywords:** Myasthenia Gravis, MG-ADL, Clinical Phenotyping, Machine Learning, Clinical Trial Endpoints, Trajectory Analysis, MGTX trial

## Abstract

**Objective:** Myasthenia gravis clinical trials increasingly rely on the MG Activities of Daily Living total score to assess disease severity and treatment response. However, aggregate scores may obscure clinically meaningful changes in individual symptom domains. To determine whether item-level MG-Activities of Daily Living profiles identify distinct clinical phenotypes and whether longitudinal phenotype transitions reveal disease changes inadequately captured by aggregate scores.

**Methods:** MG-Activities of Daily Living evaluations from seven clinical trials and observational studies (5,259 observations were analyzed using principal component analysis and K-medoids clustering with Cityblock distance. Longitudinal phenotype transitions were evaluated in the three longitudinal cohorts and compared with changes in total scores.

**Results:** Despite low median MG-ADL scores (2.0–3.0), substantial intercohort symptom heterogeneity was observed. Clustering identified five phenotypes: mild ocular (23.7%), severe ocular-dominant (23.5%), bulbar-respiratory dominant (22.6%), severe generalized (16.0%), and proximal limb-axial weakness (14.3%). Longitudinal phenotype instability occurred in 38–49% of Explore participants, 47–49% of Promise participants, and 20–25% of MGTX participants. Importantly, clinically meaningful phenotype transitions, including shifts from ocular to bulbar predominance, frequently occurred with changes of only one point in total MG-ADL scores. Conversely, substantial total score fluctuations could occur without a change in phenotype.

**Interpretation:** MG-ADL total scores incompletely characterize disease heterogeneity and longitudinal symptom evolution. Incorporating item-level phenotyping into clinical trials and routine care may improve interpretation of treatment response, identify clinically important deterioration overlooked by aggregate scores, and facilitate more precise assessment of disease trajectories.

## Introduction

Neurological drug development continues to rely heavily on clinical rating scales developed decades ago, many of which remain accepted standards for evaluating treatment efficacy. However, these instruments have important limitations, including floor and ceiling effects and the potential loss of clinically meaningful information when multiple functional domains are combined into a single composite score.(1–3) Multidomain ordinal scales are also widely used as primary trial outcomes in diseases such as amyotrophic lateral sclerosis, Parkinson disease, Friedreich ataxia, Alzheimer disease, Rett syndrome, and select non-neurological conditions. These instruments combine graded impairment across several functional domains into a single score. This is practical and clinically interpretable, but the total can conceal important differences: patients with the same score may have very different patterns of disability, disease biology, prognosis, and treatment response.

Myasthenia gravis (MG) is a rare, clinically and biologically heterogeneous autoimmune disorder of the neuromuscular junction characterized by fluctuating skeletal muscle weakness. (4) Variation in symptom distribution, severity, disease course, and treatment response, both among patients and within individuals over time, presents considerable challenges for clinical trial design and personalized treatment assessment. The MG Activities of Daily Living (MG-ADL) scale has become a principal efficacy endpoint in MG clinical trials and has supported regulatory evaluations by the US Food and Drug Administration and European Medicines Agency.(5) The instrument assesses eight functional domains encompassing ocular symptoms (ptosis and diplopia), bulbar function (speech, chewing, and swallowing), respiratory function, and limb function (brushing teeth or combing hair and rising from a chair).(6) Despite its widespread acceptance, concerns remain regarding its ability to adequately capture the heterogeneity of disease manifestations and changes in individual patients.(7–9) In particular, identical total scores may represent markedly different symptom profiles, while equivalent changes in aggregate scores may reflect improvement or deterioration in different functional domains.

In this study, we integrate item-level MG-ADL data across multiple clinical trial and research populations to address two fundamental questions. First, can distinct clinical phenotypes be identified from patterns of individual MG-ADL items that are obscured by aggregate scores? Second, how do these phenotypes change over time, and what are the implications of these transitions for interpreting clinical improvement, treatment response, and trial endpoints? By examining symptom profiles and their longitudinal evolution, we seek to determine whether item-level analyses provide clinically meaningful information beyond conventional total-score assessments.

More broadly, this approach provides a foundation for phenotype informed interpretation of clinical outcomes and the development of computational models that capture individual disease trajectories. Incorporating the distribution and evolution of functional impairments, rather than relying exclusively on aggregate scores, may improve clinical trial endpoint interpretation, support individualized treatment assessment, and inform the development of more sensitive outcome measures. These principles may extend beyond MG to other neurological and systemic disorders evaluated using multidomain ordinal scales.

## Methods

We assembled a pooled dataset from the clinical studies and trials: Muscle Study Group trial of mycophenolate versus prednisone,(10) the MGTX Study,(11) BeatMG,(12) RINOMAX, (13), ADAPT-teleMG (NCT 05917184), Promise,(14) and the EXPLORE-2 MGNet natural history study (NCT 03792659) (Table 1). Each record included scores for eight MG-ADL items, each rated on a 0–3 ordinal scale: ocular function (eyelid droop and diplopia/vision), bulbar function (talking, chewing, and swallowing), respiratory function (breathing), and axial/limb function (rising from a chair and brushing hair/teeth). Completed assessments were classified as:

- Mild/Remission: Total MG-ADL Score *≤* 2 (*n* = 2,821).
- Symptomatic Cohort: Total MG-ADL Score > 2 (*n* = 2,438).

**Table 1.** Clinical Studies in Analysis.

| **Study Name/NCT** | **Description** | **Subject Number** | **Discrete MG-ADL Measures** |
| --- | --- | --- | --- |
| Muscle Study Group<br>00683969 | 12 week randomized,<br>24 week open label<br>extension, Phase 3 | 80 | 551 |
| MGTX<br>00294658 | 3-year phase 3 trial | 111 | 1,907 |
| BeatMG<br>02110706 | 1-year phase 2 trial | 43 | 705 |
| Rinomax<br>02950155 | 16 week randomized<br>trial | 47 | 328 |
| Adapt-TeleMG<br>05917184 | 2 visit virtual<br>assessments video<br>recordings | 51 | 104 |
| Explore | 2-year natural history<br>study | 302 | 1,176 |
| 03792659 |  |  |  |
| Promise<br>03490539 | 2-3 year<br>observational study | 174 | 488 |

To evaluate the clinical trial cohorts can be considered comparable in the composition of individual severity scores, we first compared aggregate total-score distributions across cohorts using medians, interquartile ranges (IQRs), and boxplots. We then used Mahalanobis distance in the eight-dimensional item space to determine whether individual study cohorts differed systematically from the pooled multi-study population:

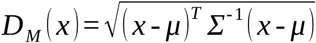

where *x* represents an individual patient vector, *μ* is the multi-study population mean vector, and *Σ* is the covariance matrix. A profile cutoff threshold corresponding to the 99th percentile of the chi-square distribution with 8 degrees of freedom (*D_M_* = 4.48) was established to identify systemic population-level outliers.

Principal component analysis (PCA) was applied to the standardized matrix of eight item scores to examine correlations, redundancy, and grouping among items.(15) We assessed the variance explained by successive components and generated PC1-versus-PC2 loading biplots to visualize the relationships among ocular, bulbar, respiratory, and limb/axial features. Phenotypic clustering was performed strictly on the symptomatic evaluation subset (Total Score > 2; *n* = 2,438) to prevent floor-effect distortion from asymptomatic/mild visits.

We employed K-medoids clustering (PAM - Partitioning Around Medoids). Unlike K-means, K-medoids selects actual data points (exemplars) as cluster centers, making it robust to discrete ordinal scores and outlier evaluations.(16). Distance between patient sub-item profiles *u* and *v* was computed using the Cityblock (Manhattan/L1) distance (17):

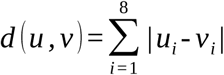

The *L* 1 metric is strictly aligned with the natural additive structure of ordinal clinical rating scales like the MG-ADL.

The optimal number of clusters *k* was determined by evaluating the Elbow Method on the total Within-Cluster Sum of Squares (WCSS) across *k* = 1 through *k* = 8. Models at *k* = 3 and *k* = 5 were evaluated against clinical interpretability and functional domain coverage.

To evaluate how patient phenotypes evolve over time, longitudinal tracking was performed on cohort subsets with at least four consecutive visits: the Explore observational cohort (*n* = 173), the Promise observational cohort (n=301) and the MGTX clinical trial cohort (n=111).

For each patient *j* evaluated across *T* visits with phenotypic assignments *C _j_* =(*c*_1_ *, c*_2_ *,…, c_T_*), trajectory instability was quantified via the Total Variation (TV) of class transitions:

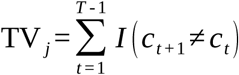

where *I* (*⋅*) is the indicator function yielding 1 if a phenotypic boundary shift occurred between consecutive visits, and 0 otherwise. Trajectory instability was quantified via the Total Variation (TV_j) of class transitions across visits. To focus strictly on clinically meaningful, high-frequency phenotypic switching rather than isolated score noise, a trajectory was classified as highly unstable if TV_j >= 0.5, indicating that the patient transitioned between distinct phenotypic categories on average at least once every two visits.

To evaluate the agreement between qualitative phenotypic class transitions and the standard Minimal Clinically Important Difference (MCID, defined as a *≥±* 2-point total score change), we calculated:

1. The normalized frequency of class changes per visit (TV_norm_).
2. The normalized frequency of total score shifts *≥* 2 points (MCID_norm_).

Pearson correlation coefficients were used to assess the degree of correspondence between MCID-based total-score changes and transitions between item-level phenotypes.

To determine whether phenotype transitions reflected genuine redistribution of clinical scores rather than measurement noise, we compared the total variation of the unweighted MG-ADL score with that of domain-weighted profiles across clinical classes. For encounters with substantial changes in the total or weighted score between consecutive visits, we calculated Pearson correlations between the two measures. We also compared the longitudinal vectors of the eight raw MG-ADL items with the weighted MG-ADL vector for individual patients.

Representative trajectories are provided (Supplement).

## Results

### Demographic Heterogeneity Across Study Cohorts

We assembled a pooled dataset of 5,259 MG-ADL assessments from seven clinical trials and natural history studies, representing 808 patients (Table 1). In addition to differences in clinical severity, the seven studies differed substantially in demographic composition, particularly age and gender distributions (Supplemental Figs A1-A7). Entrance criteria for nearly all studies was positive serology of AChR antibodies. Only, the EXPLORE study entered 39 MuSK antibody-positive and 38 seronegative patients or no listing of antibody status.

Sex distributions varied across cohorts. BeatMG (45% women; Fig A2) and ADAPT (51% women; Fig A3) were relatively balanced, whereas MGTX enrolled a predominantly women (71% Figure A4). By contrast, men compromised the majority in Explore (57%, Fig A1); PROMISE (61%; Figure A7), and Rinomax (70%; Fig A6). Age distributions reflected differences inclusion criteria of the studies. MGTX enrolled a younger population, concentrated between 18 and 50 years of age and peaking at 20–35 years (Fig A4), consistent with its inclusion of patients younger than 65 years. MSG and ADAPT showed a bimodal distribution, with both early-onset (30–40 years) and late-onset (65–75 years) groups (Figs A3 and A5). In contrast the Explore,

PROMISE, and Rinomax were skewed toward older patients, with peaks between 65 and 75 years and few participants younger than 40 years (Figs A1, A6, and A7). These studies did not have an upper age restriction for participation.

### Cross-Study MG-ADL Distributions and Cohort Heterogeneity

After excluding approximately 11% of incomplete records (all from the Promise study which allowed submission of either aggregate or individual scores), the pooled dataset comprised 5,259 MG-ADL assessments. Because the studies used different conventions for variables such as sex, age, and item order, all datasets were harmonized within a common coding framework. As shown in the combined macro/micro analysis (Table A11), total-score distributions were broadly similar: medians ranged from 1.0 to 4.0 of 24, and means ranged from 2.40 to 4.26. The IQRs overlapped substantially across all seven cohorts (Fig 1A), and Mahalanobis distance analysis did not identify systematic study-level divergence from the pooled cohort centroid (Fig 1B).

**Figure 1.**
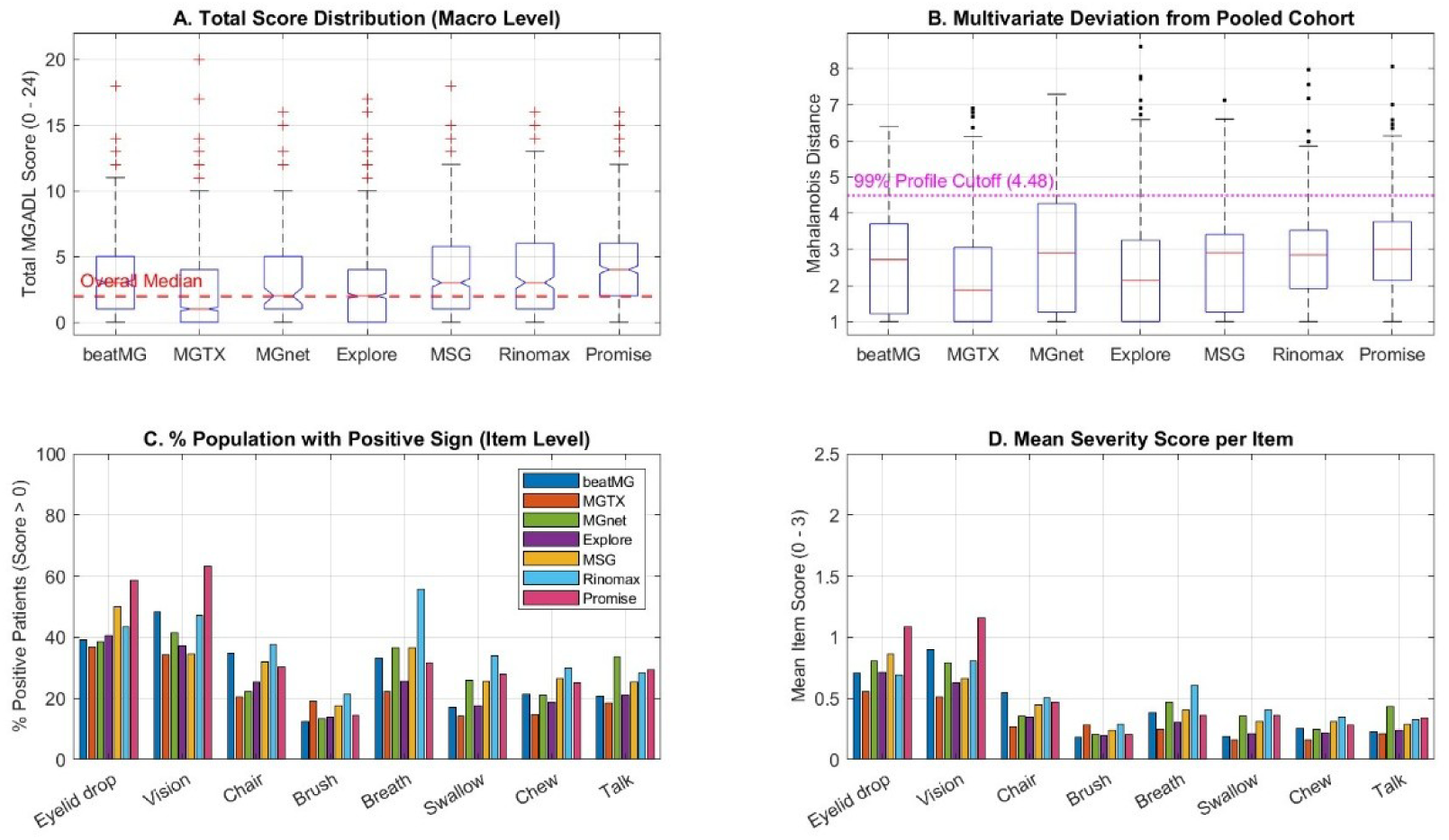
Overview comparing the clinical studies. Mean MG severity, as measured by the MG-ADL, across cohorts and across specific muscle groups. Rinomax (Study R) was an outlier, with greater impairment across the individual items. Across the clinical studies, total MG-ADL scores had low baseline medians (2.0–3.0) and broadly overlapping boxplot distributions. Item-level incidence, however, showed that Study R (Rinomax, N = 328) included a more symptomatic cohort. Positive findings were more frequent across almost all eight items, particularly for breathing (>55 with shortness of breath, compared with 22–37 in the other cohorts) and bulbar function (swallowing, chewing, and speech). Thus, although aggregation and floor effects concealed this difference in the total-score boxplots, item-level prevalence showed a broader multidomain disease burden in Study R.

Sub-item profiling revealed profound underlying phenotypic heterogeneity among trial cohorts (Fig 1C, 1D). In six of the seven studies (BeatMG, MGTX, ADAPT, Explore, MSG, and PROMISE), disease burden was heavily ocular-dominant, with eyelid drooping (36.8%–58.2%) and double vision (35.2%–63.3%) representing the most prevalent symptoms. In contrast, Rinomax (*N* = 328) enrolled a systematically distinct cohort characterized by severe multi-domain involvement: 55.8% of Rinomax patients presented with positive respiratory signs (Breath), compared to 22.2%–36.8% in other studies. Rinomax also demonstrated marked elevations in bulbar prevalence (33.8% Swallow, 29.9% Chew, 28.0% Talk).

The addition of the PROMISE observational cohort (*N* = 488 valid evaluations) highlights a distinct baseline profile relative to established trial cohorts (Fig 1, Table A11). While randomized clinical trial cohorts (e.g., MGTX, BeatMG) and longitudinal registries (Explore) include substantial proportions of mildly symptomatic or asymptomatic follow-up evaluations (total score *≤* 2), the PROMISE cohort demonstrated a higher baseline median score (4.0) and a minimal proportion of asymptomatic evaluations (Fig 1A, Table A11). At the item level, PROMISE exhibited the highest ocular domain severity in the multi-study population, with positive sign incidence reaching 58.2% for eyelid drop and 63.3% for diplopia/vision impairment, alongside higher mean item severity scores (> 1.0; Fig 1C, 1D, Table A11).

These findings confirm that aggregate total scores obscure critical clinical heterogeneity between trial populations. Variations in sub-item profiles reflect study design differences: Rinomax required a QMG score *≥* 6 (selecting for broader systemic/respiratory involvement), Explore captured MuSK MG patients with known bulbar predominance, and PROMISE enrolled an inception cohort of newly presenting patients evaluated prior to long-term therapeutic control.

### Dimensionality Reduction and Cluster Optimization

To uncouple sub-item interactions, PCA was performed on all 5,889 evaluations (*N* = 5,259 valid assessments). Five principal components accounted for *∼* 80 % of total variance. Biplot analysis demonstrated distinct spatial groupings among ocular items (Eyelid Drop, Vision) and bulbar items (Chew, Swallow, Talk), while axial and limb items (Chair, Brush, Breath) projected along independent vectors (Fig 2).

**Figure 2.**
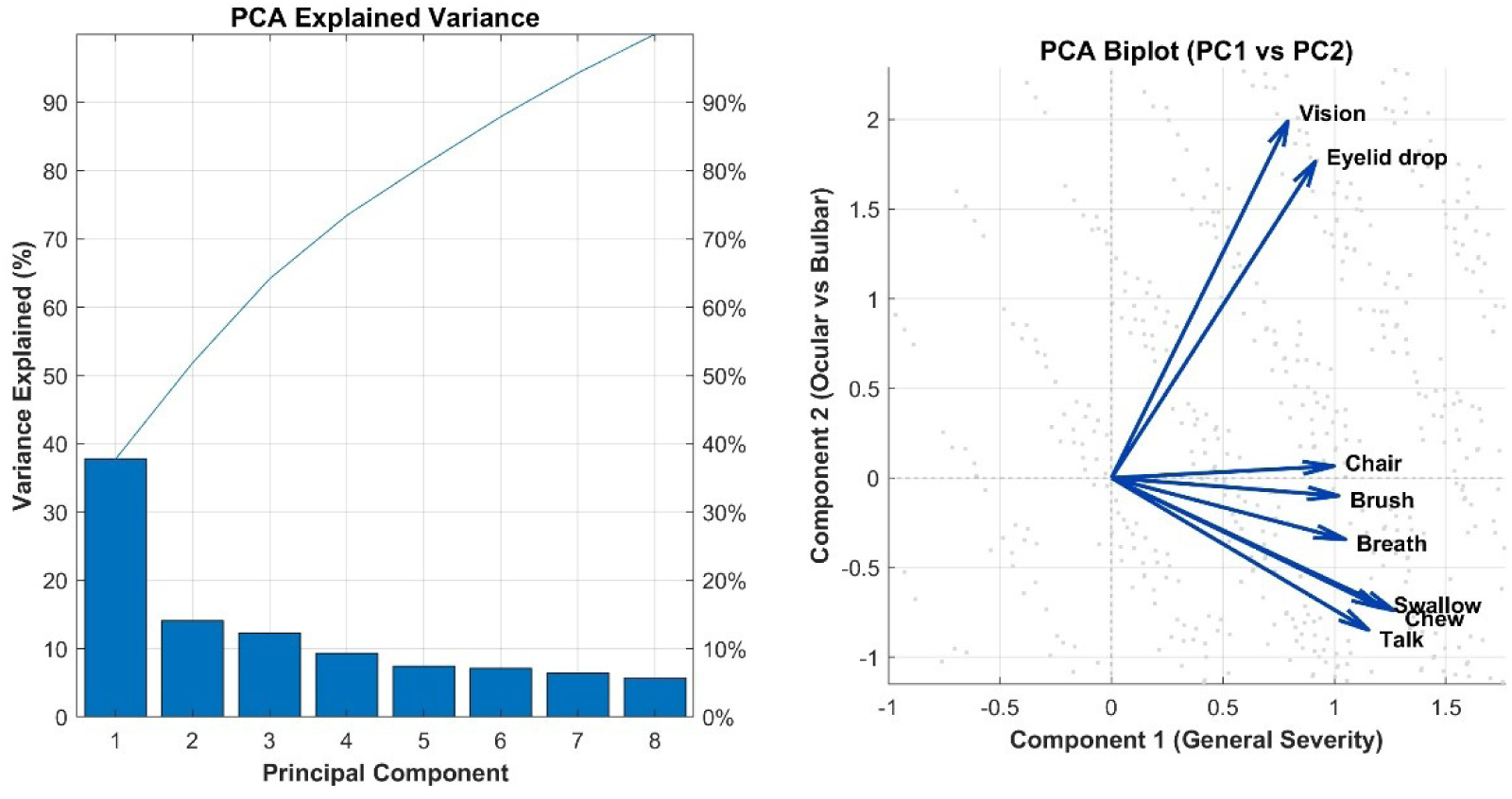
Principal component analysis of the pooled MG-ADL dataset. Including 2,821 assessments in the mild/remission group (total score <=2) and 2,438 in the symptomatic group (total score >2), five principal components explained approximately 80% of the variance. Ocular items (vision and eyelid droop) and bulbar items (chewing, swallowing, and talking) formed distinct groups.

Restricting phenotypic classification to symptomatic assessments (total MG-ADL > 2; *n* = 2,438), K-medoids clustering with Cityblock distance was evaluated across *k* = 1 to *k* = 8 clusters. The total Within-Cluster Sum of Squares (WCSS) elbow curve demonstrated a major inflection between *k* = 3 (*∼* 5,900) and *k* = 5 (*∼* 4,900), beyond which variance compression entered a linear plateau (Fig 3). While a 3-cluster model captured broad Ocular-Dominant (Phenotype A: 42.5%, *n* = 1,036), Bulbar-Dominant (Phenotype B: 33.1%, *n* = 808), and Generalized (Phenotype C: 24.4%, *n* = 594) classes (Figs A13 & A14), it failed to separate axial/limb and respiratory involvement from general severity. Consequently, the 5-cluster model was selected for granular phenotypic mapping (Fig 4).

**Figure 3.**
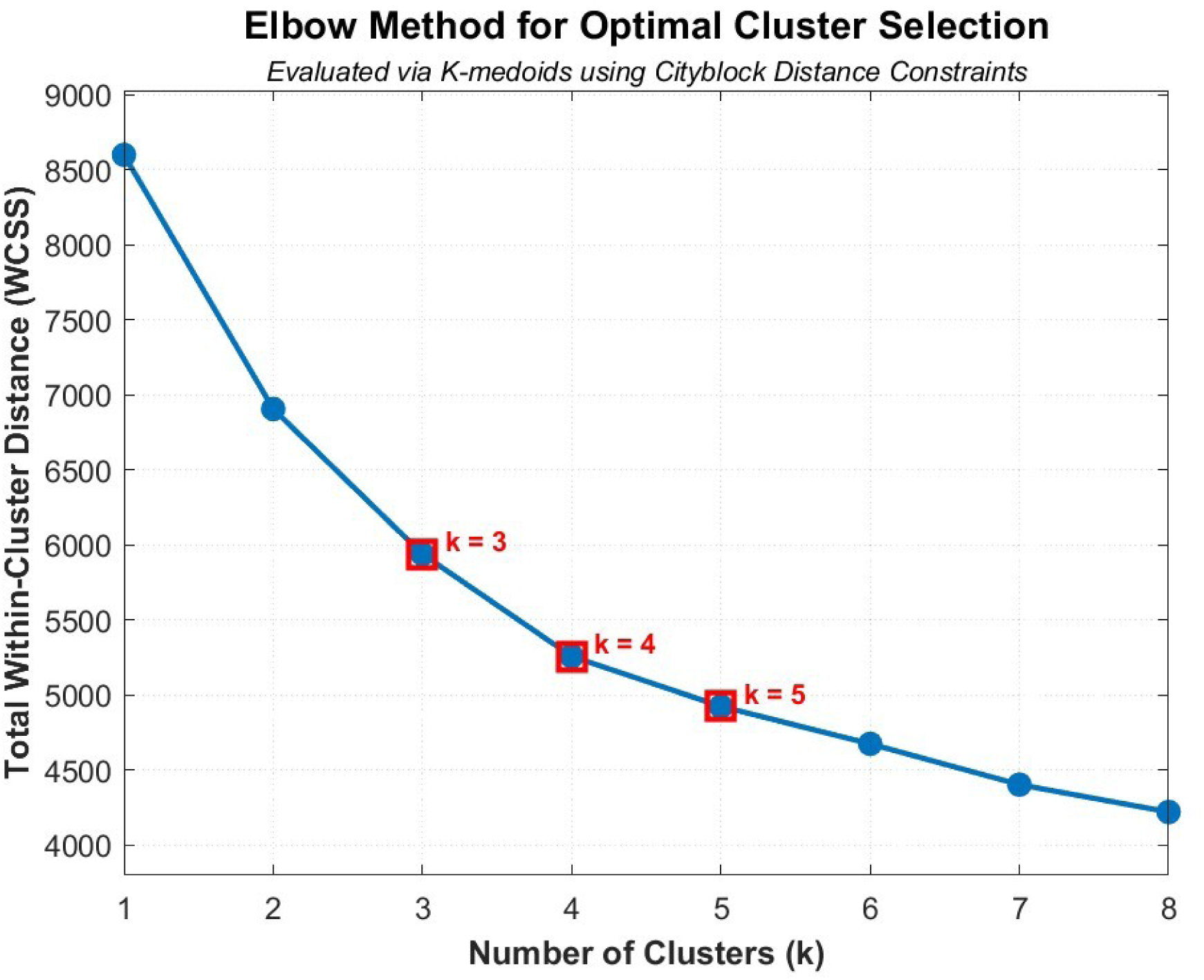
Five clusters using elbow method. The elbow method was used to assess the number of clusters for classifying symptomatic MG-ADL profiles and increased the number of clusters from three to five reduced the within-cluster sum of squares (WCSS) from approximately 5,600 to 4,600. At k=5, the curve reached the turning point before further variance reduction became approximately linear.

**Figure 4.**
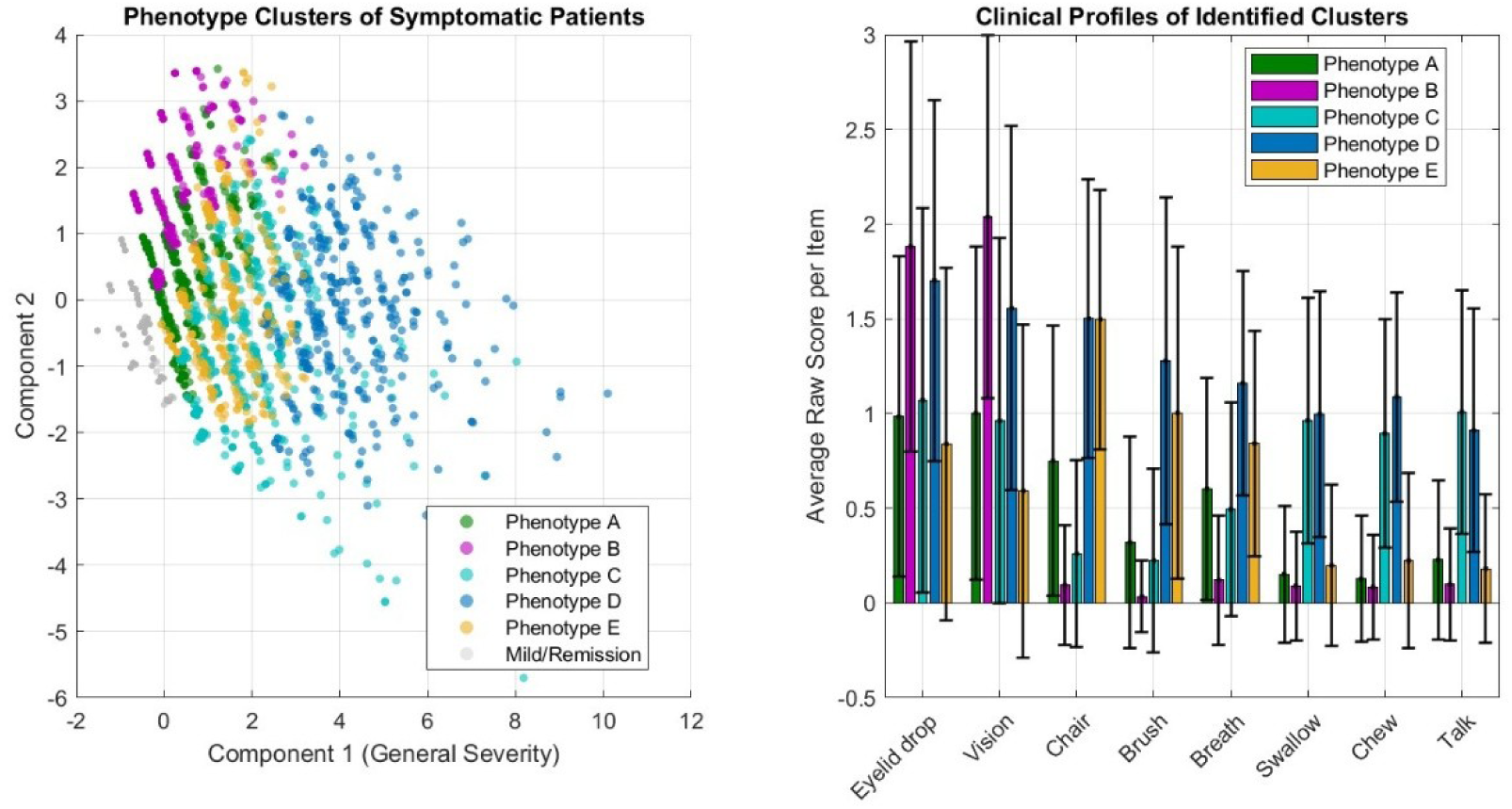
Phenotypic Classification. The five clusters corresponded to Phenotype A. (*n* = 577,23.7 %): Mild Ocular / Baseline Symptomatic (green), Phenotype B (*n* = 574,23.5 %): Severe Ocular-Dominant (purple), Phenotype C (*n* = 550,22.6 %): Bulbar & Respiratory-Dominant (cyan), Phenotype D (*n* = 389,16.0 %): Severe Generalized / Pan-Domain (dark blue), and Phenotype E (*n* = 348,14.3 %): Proximal Limb & Axial Weakness (orange)

### Identification of Five Discrete MG-ADL Phenotypes

Unsupervised K-medoids clustering (*k* = 5) of symptomatic evaluations (Total Score > 2; *n* = 2,438) stratified patients into five discrete anatomical and functional phenotypes (Fig 4), ordered by cluster size:

- **Phenotype A (Mild Ocular / Baseline Symptomatic; Green):** Characterized primarily by mild ocular involvement (Eyelid Drop mean *≈* 0.98, Vision mean *≈* 1.04) and near-zero bulbar, respiratory, or axial/limb weakness, clustering near the mild symptomatic boundary on PC1.
- **Phenotype B (Severe Ocular-Dominant; Purple):** Characterized by high ocular burden (Eyelid Drop mean *≈* 1.83, Vision mean *≈* 2.04) with minimal bulbar involvement, projecting into the positive territory of the ocular axis (PC2).
- **Phenotype C (Bulbar & Respiratory-Dominant; Cyan):** Defined by predominant bulbar involvement (Talking mean *≈* 1.02, Swallowing mean *≈* 0.94, Chewing mean *≈* 0.91) accompanied by elevated respiratory compromise (Breath mean *≈* 0.48), projecting into the negative bulbar axis of PC2.
- **Phenotype D (Severe Generalized / Pan-Domain; Dark Blue):** Displays severe, multi-domain impairment across all functional items, notably combining high axial/limb scores (Chair mean *≈* 1.48, Brush mean *≈* 1.28), bulbar weakness (Swallow *≈* 1.00, Chew *≈* 1.08, Talk *≈* 0.91), and respiratory compromise, projecting furthest along PC1 (General Severity).
- **Phenotype E (Proximal Limb & Axial Weakness; Orange):** Characterized by prominent proximal fatigue and axial impairment (Chair mean *≈* 1.50, Brush mean *≈* 1.00, Breath mean *≈* 0.85) with moderate bulbar and ocular involvement.

**Figure 5.**
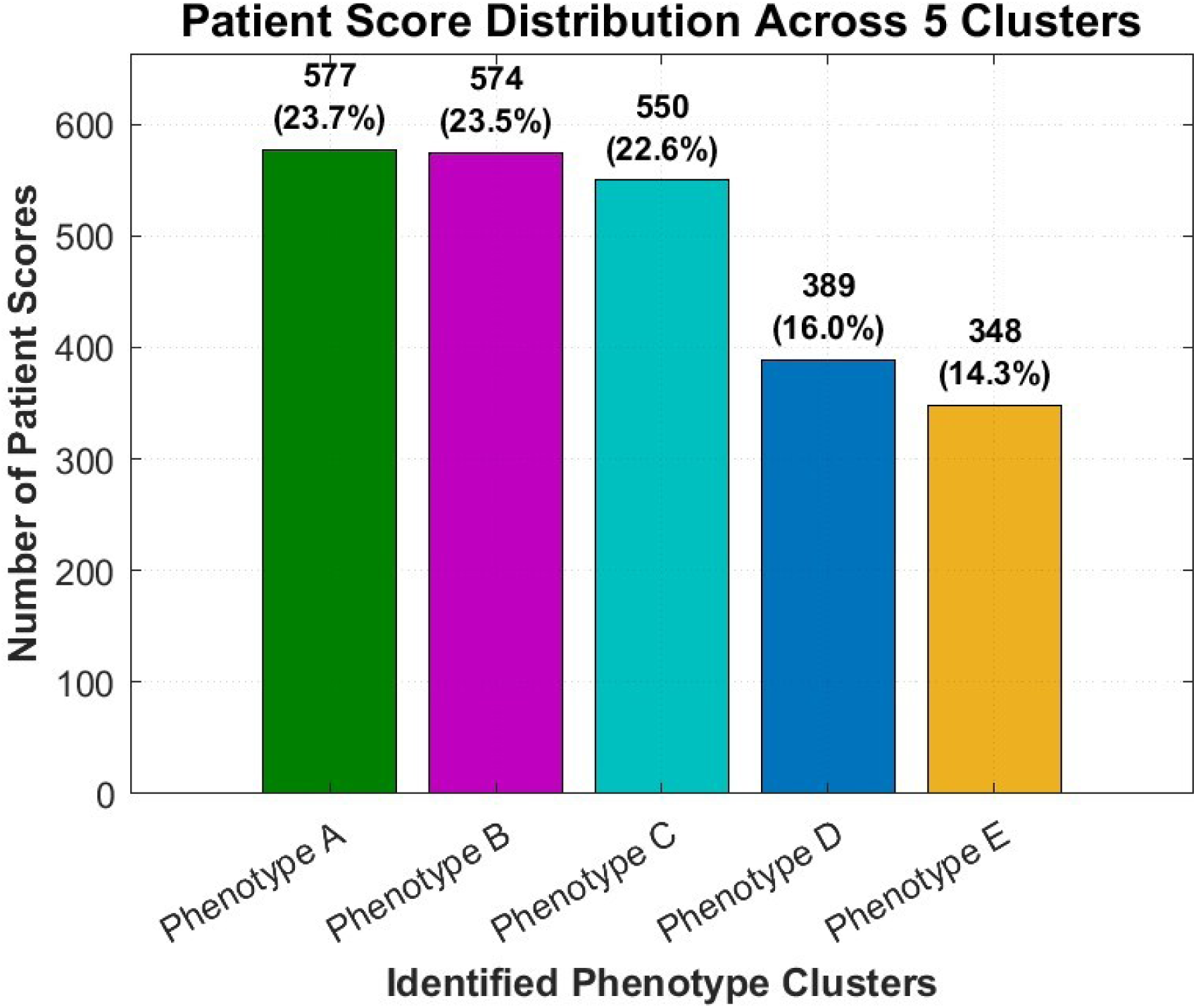
Distribution of symptomatic assessments across the five clusters shown in. Figure 4.

### Longitudinal Phenotypic Instability via Total Variation of Class Transitions

To evaluate the longitudinal trajectory of patient phenotypes, we examined patients across both observational natural history settings (Explore, PROMISE) and controlled interventional trial environments (MGTX). We defined phenotypic instability using the Total Variation (*TV*) of class transitions along a patient’s timeline of consecutive visits. A trajectory was classified as unstable if the patient transitioned between different phenotypic cleuthsters at least once across their visit timeline (*T V _j_* > 0).

#### Dynamics in real-world observational cohorts

In the Explore cohort (*n* = 181 with *≥* 4 visits), 38% (3-cluster model Figure 6 Panel A) to to 39% (5-cluster model Figure 6 Panel B) of patients exhibited longitudinal phenotypic instability (*T V _j_* > 0.5) in class assignment, with 33% representing persistently asymptomatic states (total score *≤* 2) and *≤* 3 % maintaining a constant symptomatic phenotype (Fig 6). The PROMISE cohort longitudinal tracking demonstrated even higher trajectory dynamics (Fig A11). Persistent single-phenotype trajectories were virtually absent (*≤* 2 %), while persistent asymptomatic evaluations accounted for only **9%** of the follow-up assessments. Crucially, **47%** (3-cluster model-Figure A11 panel A) to **49%** (5-cluster model – Figure A11 Panel B) of PROMISE patients exhibited longitudinal phenotypic instability (*T V _j_* > 0.5), dynamically crossing domain-specific cluster boundaries over time.

**Figure 6.**
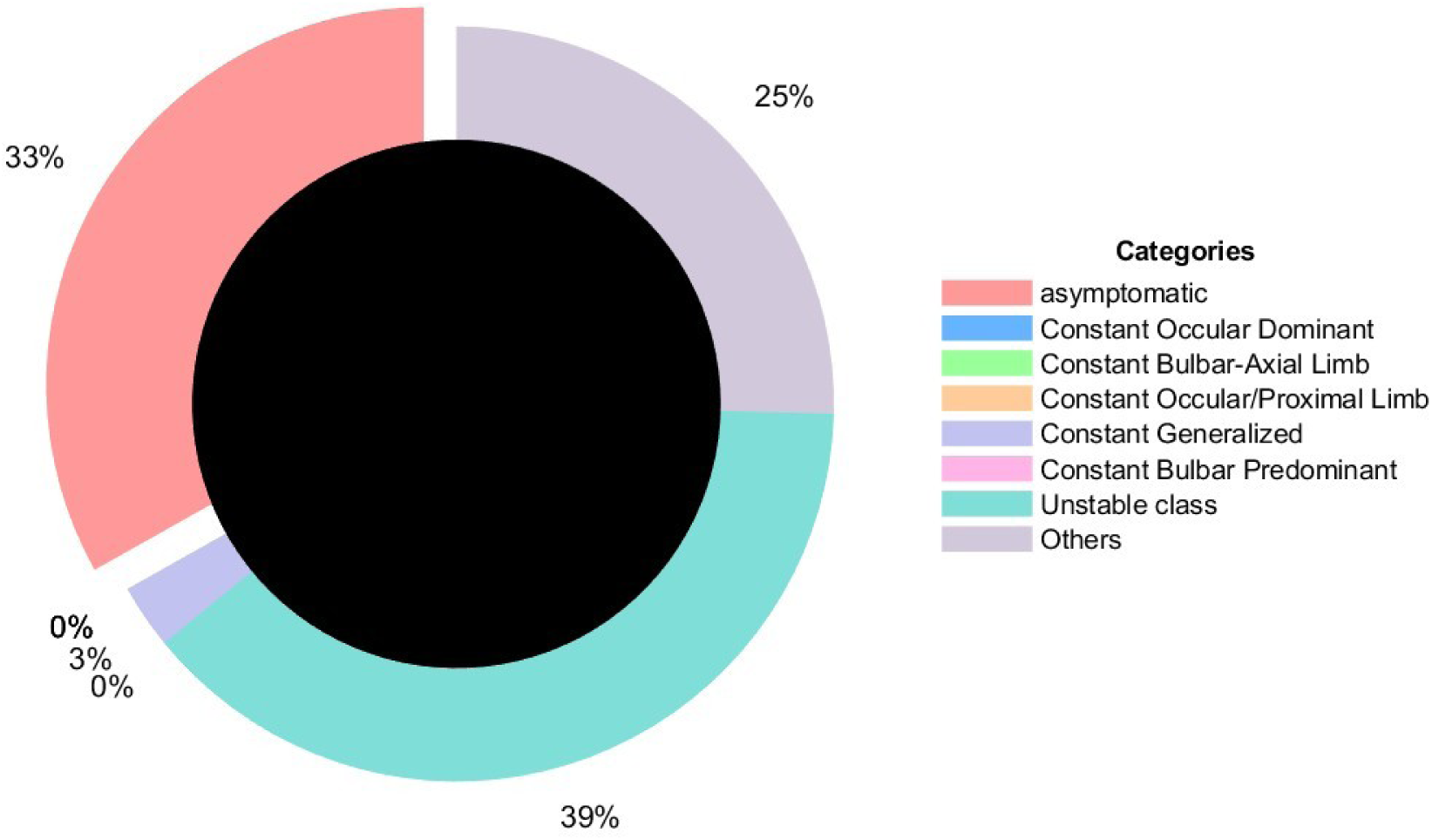
Stability of Clinical Phenotypes. Stability of patient phenotypes over at least four visits in Explore cohort, using the three-cluster model (A) and the five-cluster model (B). Approximately 38%–39% of patients had unstable longitudinal MG-ADL phenotypes, defined as a change in cluster assignment at least once over three visits. The analysis was restricted to patients with at least four visits; 181 of 301 patients in Explore met this criterion.

#### Dynamics in a controlled interventional trial (MGTX)

Using the same approach for the MGTX trial, which used a structured treatment protocol (Fig 7), we found a persistently asymptomatic group with 20% of longitudinal trajectories remaining asymptomatic throughout follow-up, while persistent phenotype trajectories were exceptionally rare. Only 20% of patients in the three-cluster model (Figure 7 Panel A) and 25% in the five-cluster model (Figure 7 Panel B) showed unstable longitudinal MG-ADL patterns. Phenotypic instability was less frequent in MGTX (20%–25%) than in Explore (38%–39%) or PROMISE (47%–49%). This difference is consistent with greater stabilization of domain-specific symptoms under standardized treatment protocols and continuous immunosuppressive management in a controlled trial setting.

**Figure 7.**
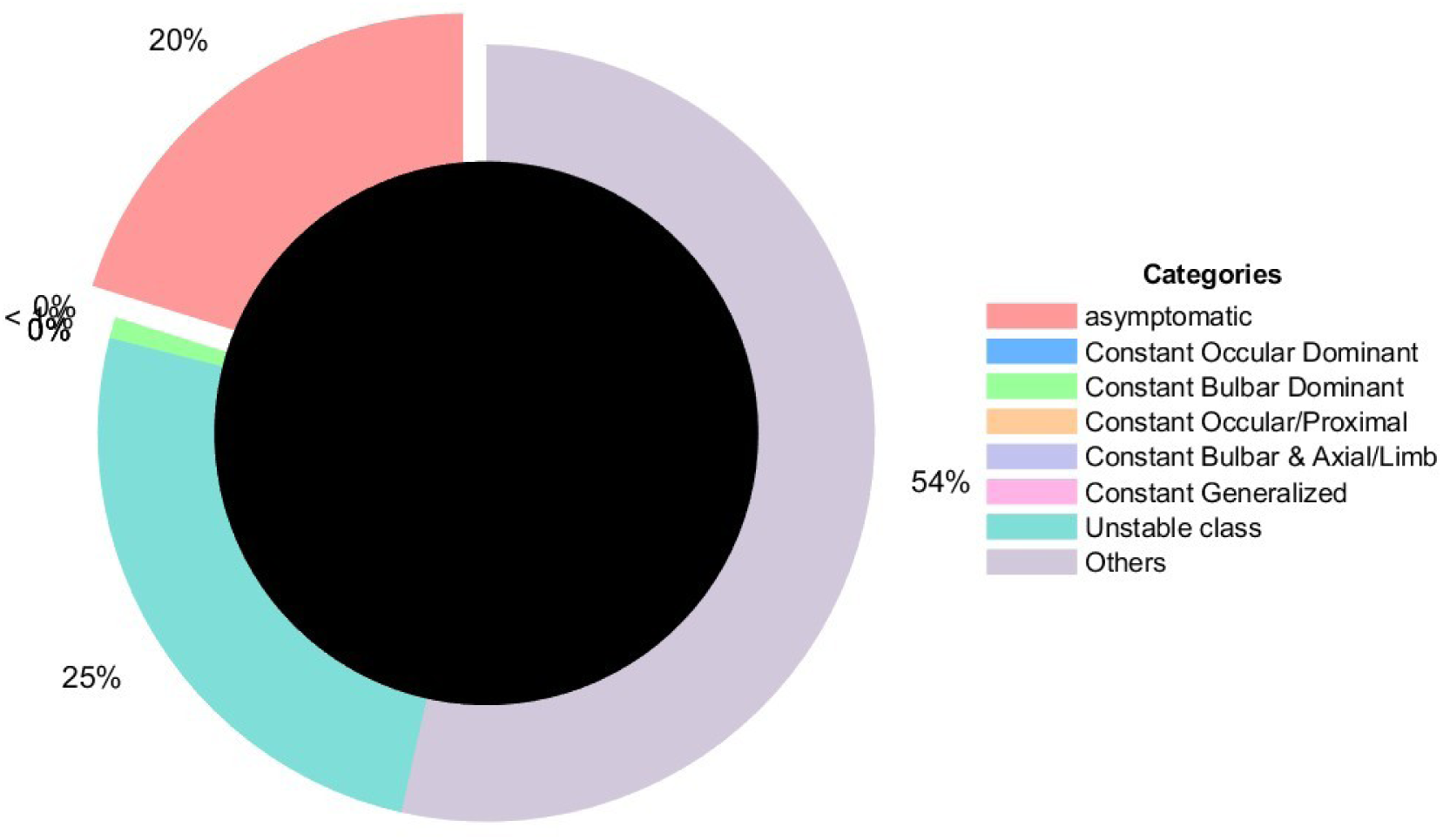
Stability of patient phenotypes in MGTX. Using the three-cluster model (A) and the five-cluster model (B), approximately 20%–25% of patients had unstable longitudinal MG-ADL phenotypes, defined as a change in cluster assignment at least once over three visits. The analysis was restricted to patients with at least four visits, which represented a large majority of the cohort (Figure A9). Instability was less frequent than in the Explore cohort.

### Re-Evaluating the MCID “2-Point Rule” Against Class Transitions

Finally, we examined the relationship between the total variation of phenotypic class transitions and the standard MCID, universally defined as a *≥±* 2-point shift in total MG-ADL score.

Two-dimensional scatter plots comparing normalized class transition frequency against normalized MCID total score shifts revealed significant structural decoupling: there is no correlation between the total variation of phenotype changes and the MCID for patients exhibiting non-zero trajectory variation (*TV ≥* 0.5; see Supplement Table 2).

This lack of correspondence appeared in two clinically distinct patterns. 1) Score Shifts Without Class Transitions: Multiple patients demonstrated frequent *≥* 2-point total score fluctuations (high normalized MCID changes) while maintaining a total variation of class assignment equal to zero. These patients remain bounded within a single severe phenotype (e.g., fluctuating within Phenotype E), where score variations represent depth of weakness within a domain rather than a qualitative shift in anatomical distribution. 2) Class transitions without MCID shifts: Conversely, many patients changed phenotype despite minimal variation in their total score. For example, a patient could move from ocular Phenotype A to bulbar Phenotype B with a total-score change of only 1 point.

Longitudinal trajectories were also complex beyond this categorical comparison (Fig A11). Among patients who moved through at least three phenotypes, 41% in PROMISE and 22% in Explore showed no correlation between the raw aggregate score and the domain-weighted MG-ADL score. In these patients, the conventional 2-point MCID criterion did not capture important changes in the distribution of symptoms across domains. These findings show that aggregate MCID-based assessment can miss qualitative changes in disease presentation.

These results suggest that therapeutic resistance and redistribution of primary symptoms over time may reflect sustained involvement across several domains rather than a single isolated deficit, a pattern that aggregate scoring may fail to represent.

## Discussion

In this pooled analysis of more than 5,000 MG-ADL assessments across seven clinical trials and observational cohorts, we demonstrate that despite broadly overlapping total-score distributions, there is marked heterogeneity in item-level profiles across cohorts that segregates into reproducible, anatomically distinct phenotypes. Moreover, longitudinal analyses showed that patients frequently transitioned between phenotypes over time and that these transitions were poorly related to conventional 2-point changes in MG-ADL total score. These observations suggest that aggregate scoring can obscure clinically important changes in the distribution of weakness and provide a rationale for incorporating item-level phenotypic information into the assessment of MG disease activity and treatment response. The MG-ADL scale is widely used to quantify disease severity and therapeutic response as the primary outcome in clinical trials for regulatory approval and for decisions on patients qualifying for high-cost treatments,(18, 19) yet its conventional interpretation relies almost entirely on the aggregate score. This approach assumes that equivalent total scores represent comparable clinical states and that a change of 2 points, commonly considered a MCID, adequately captures meaningful change. However, MG is characterized by marked heterogeneity in the anatomical distribution of weakness, and clinically distinct patterns of ocular, bulbar, respiratory, and limb involvement may produce similar aggregate scores.

While the MG-ADL serves as a practical, regulatory-accepted benchmark of disease burden, evaluating MG severity solely through this composite scale introduces key methodological and clinical limitations. Principal among these is the inherently subjective nature of patient self-reporting, which is susceptible to recall bias, inter-patient variance in symptom perception, and daily reporting thresholds.(20–22) Furthermore, the scale lacks complete domain coverage, omitting critical functional areas such as neck muscle strength, a primary clinical indicator of impending respiratory compromise, and distal extremity weakness. A 2-point reduction in total MG-ADL score is universally adopted as the MCID for responder analysis.(23) However, evaluating MG severity strictly through an unweighted total score treats distinct anatomical domains as mathematically equivalent, effectively masking domain-specific exacerbations through floor effects and score compression. When pooled across large multi-study cohorts, aggregate total scores exhibit remarkably low baseline medians (1.0–4.0) and overlapping interquartile ranges. Yet, granular sub-item analysis reveals profound clinical heterogeneity: while ocular symptoms dominate major cohorts such as MSG (49.9% eyelid drop) and BeatMG (48.2% diplopia), trial cohorts such as Rinomax enroll patients with dominant respiratory (55.8% shortness of breath) and bulbar impairment. The inclusion of the PROMISE cohort further highlights how trial entry timing shapes sub-item profiles: evaluating newly presenting patients (median disease duration of 5 months) prior to long-term therapeutic control yields markedly elevated ocular sub-item severity (58.2% eyelid drop, 63.3% vision) and a higher baseline median score (4.0) relative to established multi-year trial cohorts. To overcome the constraints of the aggregate score, this study leverages a consolidated dataset of 5,259 MG-ADL evaluations to establish a phenotype-aware computational framework.

Unsupervised K-medoids clustering stratified symptomatic evaluations (MG-ADL > 2; *n* = 2,438) into five discrete clinical phenotypes: Phenotype A: Mild Ocular / Baseline Symptomatic, Phenotype B: Severe Ocular-Dominant, Phenotype C: Bulbar & Respiratory-Dominant, Phenotype D: Severe Generalized / Pan-Domain, and Phenotype E: Proximal Limb & Axial Weakness. These categories are consistent with long-standing understanding of MG clinical phenotypes and the MGFA classification.(24, 25) The MGFA classification categories define ocular isolated weakness and divides generalized weakness to a bulbar predominant and more generalized weakness with grading of severity from mild to severe.

We examined the conventional 2-point MCID criterion in relation to transitions between these phenotypes. Scatter plots showed little correspondence between variation in phenotype and aggregate MCID-based changes. Patients sometimes crossed major anatomical boundaries, for example from an ocular to a bulbar phenotype, with only a 1-point change in total score, despite a clinically meaningful change in symptom distribution. Conversely, patients with severe generalized weakness could have changes of 2 points or more while remaining within the same phenotype, reflecting a change in severity rather than in the pattern of involvement. Our five phenotypes and their longitudinal behavior are consistent with, and extend, the observations of de Meel et al.(26) In their prospective registry study of 225 patients, which used a patient questionnaire of muscle group weakness at three timepoints, weakness clustered into ocular, bulbar, and neck/limb/respiratory combinations, and patients moved between point-in-time phenotypes during the course of disease. Across 5,259 assessments, our unsupervised model reproduced this anatomical organization and quantified transition dynamics, with instability in 20%–25% of patients in MGTX and 38%–49% in Explore and PROMISE. Both studies also suggest that refractory MG is associated with broader multidomain involvement; de Meel et al. reported lower remission rates for limb and respiratory deficits.

The demographic differences across the pooled cohorts are consistent with the established bimodal epidemiology of MG. Early-onset disease (before approximately 40–50 years) is more common in women, whereas late-onset disease (after approximately 50–60 years) is increasingly common in men. The apparent demographic outliers in our dataset are therefore likely to reflect recruitment criteria and protocol-specific enrichment rather than random variation. MGTX, for example, included a young, predominantly female population (71% men; peak age, 20–35 years) because it enrolled patients with AChR-positive generalized MG who were eligible for thymectomy. Explore (57% men) and PROMISE (61% men), by contrast, reflect a clinical population increasingly composed of older men with late-onset MG. These differences should be considered when trial endpoints are compared, because age and sex may influence baseline item profiles, comorbidity burden, and longitudinal disease course.

There is limited understanding for differential muscle involvement caused by MG. Ocular muscles have inherent properties that make them more susceptible to MG with neuromuscular junction structure suggesting a low safety factor for transmission failure, unique expression of the fetal AChR, providing a distinct antigenic target, and reduced levels of complement inhibitory proteins. For MuSK MG, bulbar muscles have lower levels of MuSK expression. Beyond these observations, we can only presume that the specific properties of a muscle group combined with variations in autoantibody properties over time, which have been demonstrated to occur in AChR antibody MG,(27) lead to variation in symptom expression.

Several methodological limitations of this study warrant consideration. First, several of the included clinical trials provided only cross-sectional baseline data without high-frequency longitudinal follow-ups, which restricted our trajectory instability analysis to a subset of cohorts (Explore, Promise and MGTX). Second, a potential granularity trade-off exists within the clustering methodology: while *k* = 5 offered optimal variance compression and domain separation, the discrete boundaries inherent to ordinal clinical rating scales (0–3 per item) can occasionally introduce artificial category transitions for subtle, 1-point score fluctuations near cluster edges.

These findings have implications for clinical trial design and digital health. Future work could combine patient-reported MG-ADL items with clinician-rated QMG scores and the MG-QOL15r in a multidimensional endpoint. Phenotype-aware trial measures, such as a Phenotypic Responder Index, could then capture recovery within clinically important domains, for example, resolution of bulbar or respiratory impairment, rather than relying only on an unweighted change in total score. Incorporating transitions between the five phenotypes into digital-twin models may also improve the prediction of disease exacerbations and support individualized tapering of therapies.

## Conclusion

The total MG-ADL score can conceal clinically important changes within individual anatomical domains and may not fully represent longitudinal disease dynamics in MG. In this pooled dataset of 5,259 assessments, item-level analysis with unsupervised machine learning identified five distinct clinical phenotypes. Transitions between these phenotypes did not consistently correspond to changes meeting the conventional 2-point MCID threshold, indicating that total-score endpoints alone may oversimplify treatment response. Domain-aware, phenotype-sensitive measures may therefore improve clinical trial endpoints, help identify therapeutic resistance, and support the development of personalized digital-twin approaches in neuromuscular care.(28, 29)

## Data Availability

All the data used in this study were obtained by the investigators through individual data transfer agreements from the data holders which preclude further sharing. Therefore, we cannot provide the data directly but can advise interested parties of contacts for these organizations.

## Disclaimer

During the preparation of this work, the authors used Gemini to edit and improve language readability. The underlying ideas, method, discussion and conclusions remain entirely the original work of the authors. After using this tool, the authors thoroughly reviewed, validated, and finalized the content, and take full responsibility for the integrity of the published article.

## Acknowledgements

This work was supported in part by MGNet, a member of the Rare Diseases Clinical Research Network Consortium (RDCRN; NIH U54 NS115054). Funding support for the Data Management and Coordinating Center was provided by the National Center for Advancing Translational Sciences (NCATS) and the National Institute of Neurological Disorders and Stroke (NINDS). Dr. Garbey is CEO of Care Constitution, which supported the digital tools and AI algorithms used in this work.

## MGNet Investigators Competing Interests

Dr. Mehta has received research funding from EMD Serono, Novartis, Johnson & Johnson, Takeda, and UCB. She has served on advisory boards for Amgen, J&J, and UCB and has received speaker honoraria from UCB and Amgen. She was supported by a subaward from NIH grant U54NS115054.

Dr. Habib receives research support/ honoraria from Alexion/Astra Zeneca; argenx; Bristol Myers Squibb; UCB; Immunovant; Regeneron; CabalettaBio; Amgen; Novartis; Arcellx/Gilead; Merck; Nkarta; NMDpharma; Grifols; Cartesian; Jansen/J&J; Cour pharmaceuticals; Kyverna; Vor; Vertex; MGNet (grant number:U54NS115054). He serves on DSMB for NIH/NINDS, Immunis Biomedical, and is on the trial steering committees of Kyverna, Jansen/J&J; Vertex, and Vor. He was supported by a subaward from NIH grant U54NS115054.

Dr. Juel receives consulting fees from Accordant Health and argenx. He serves on an advisory board for Alexion and Immunovant. He is supported by a subaward from NIH grant U54NS115054.

Dr. Nowak has received research support from Alexion Pharmaceuticals, argenx, Annexon Biosciences, Inc., Alira Health, Genentech Inc., Grifols, S.A., Immunovant, Inc., Momenta Pharmaceuticals, Inc. (now Janssen), Myasthenia Gravis Foundation of America (MGFA), National Institutes of Health (NIH), Ra Pharmaceuticals, Inc. (now UCB S.A.), and Viela Bio, Inc. (Horizon Therapeutics, now Amgen Inc.). He has also served as a consultant and advisor for Alexion Pharmaceuticals, argenx, Cabaletta Bio, Inc., Cour Pharmaceuticals, Immunovant, Inc., Momenta Pharmaceuticals, Inc. (now Janssen), Ra Pharmaceuticals, Inc. (now UCB S.A.), Vor Bio, and Viela Bio, Inc. (Horizon Therapeutics, now Amgen Inc.). He is supported by a subaward from NIH grant U54NS115054.

Dr. Ruzhansky receives research support from Janssen, Immunovant, COUR pharma and Argenx. She is a consultant for Amgen, Janssen, Kyverna, Alexion, Immunovant and UCB. She serves on a DSMB for Paradromics. She was supported by a subaward from NIH grant U54NS115054.

Dr. Soliven receives consulting fees from CVS Pharmacy and has served as one-time consultant for Argenx and Dianthus. She is supported by a subaward from NIH grant U54NS115054 and salary support from NIH R01 NS116262.

---

**Figure A1.**
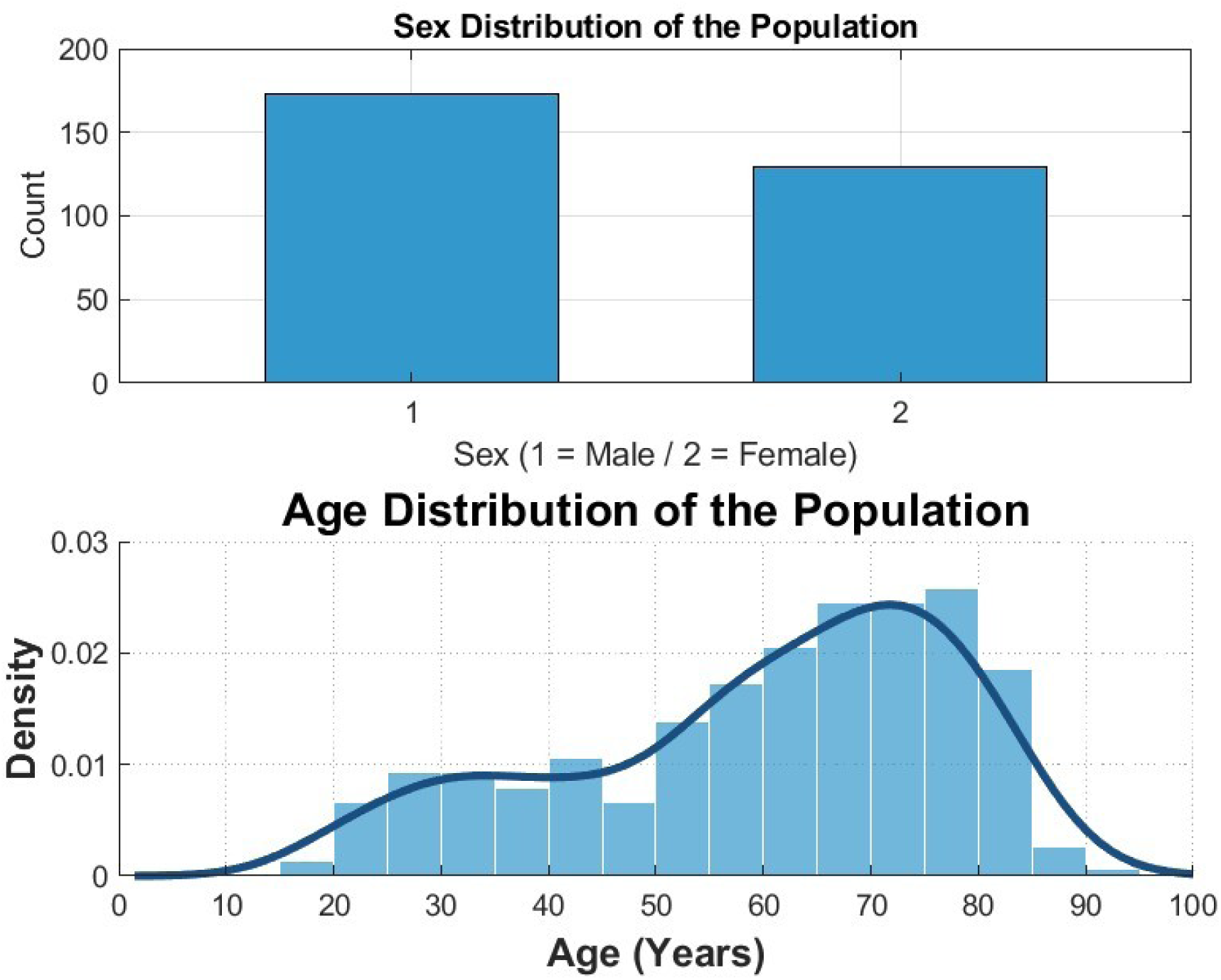
Explore Study

**Figure A2.**
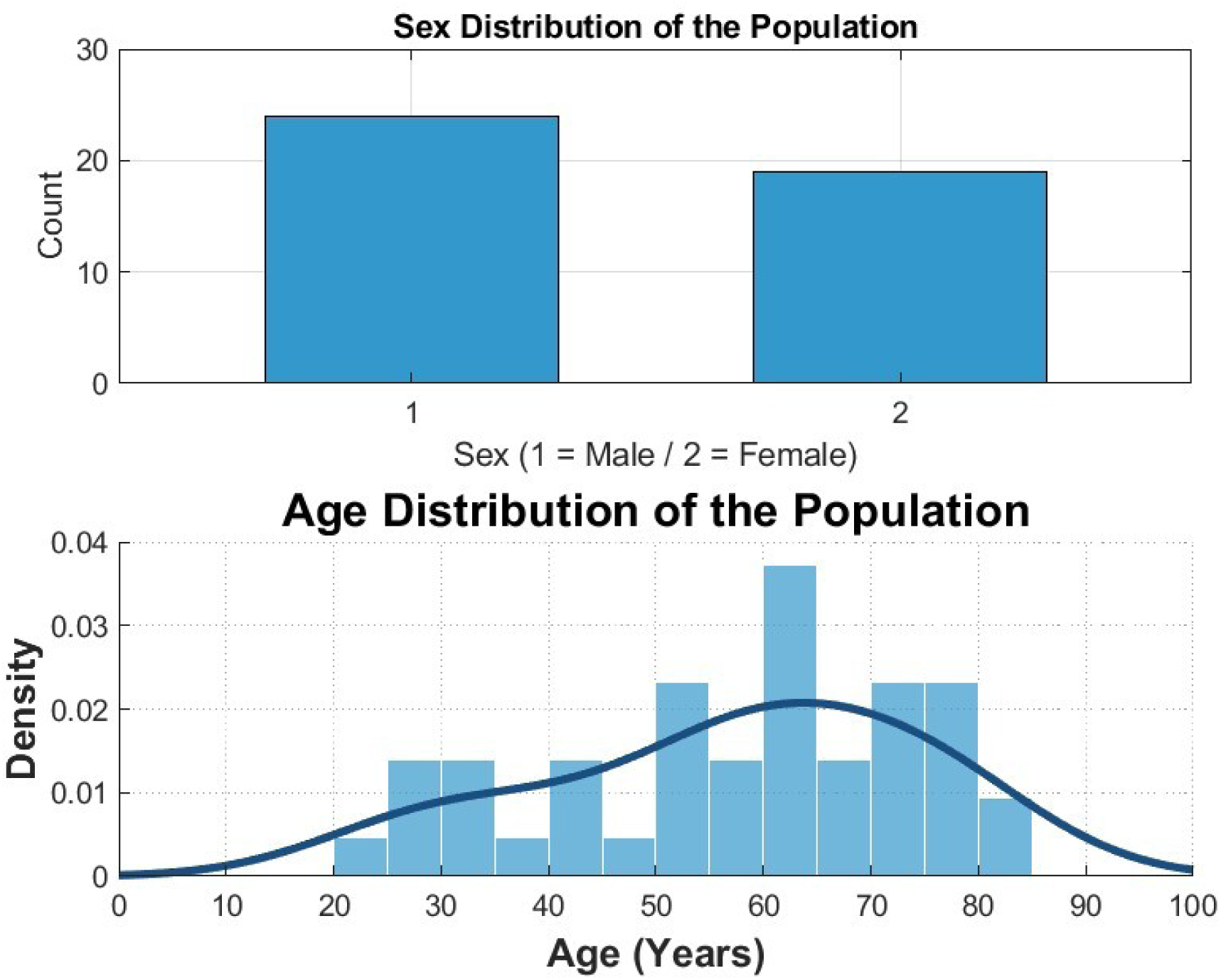
BeatMG Clinical Trial.

**Figure A3.**
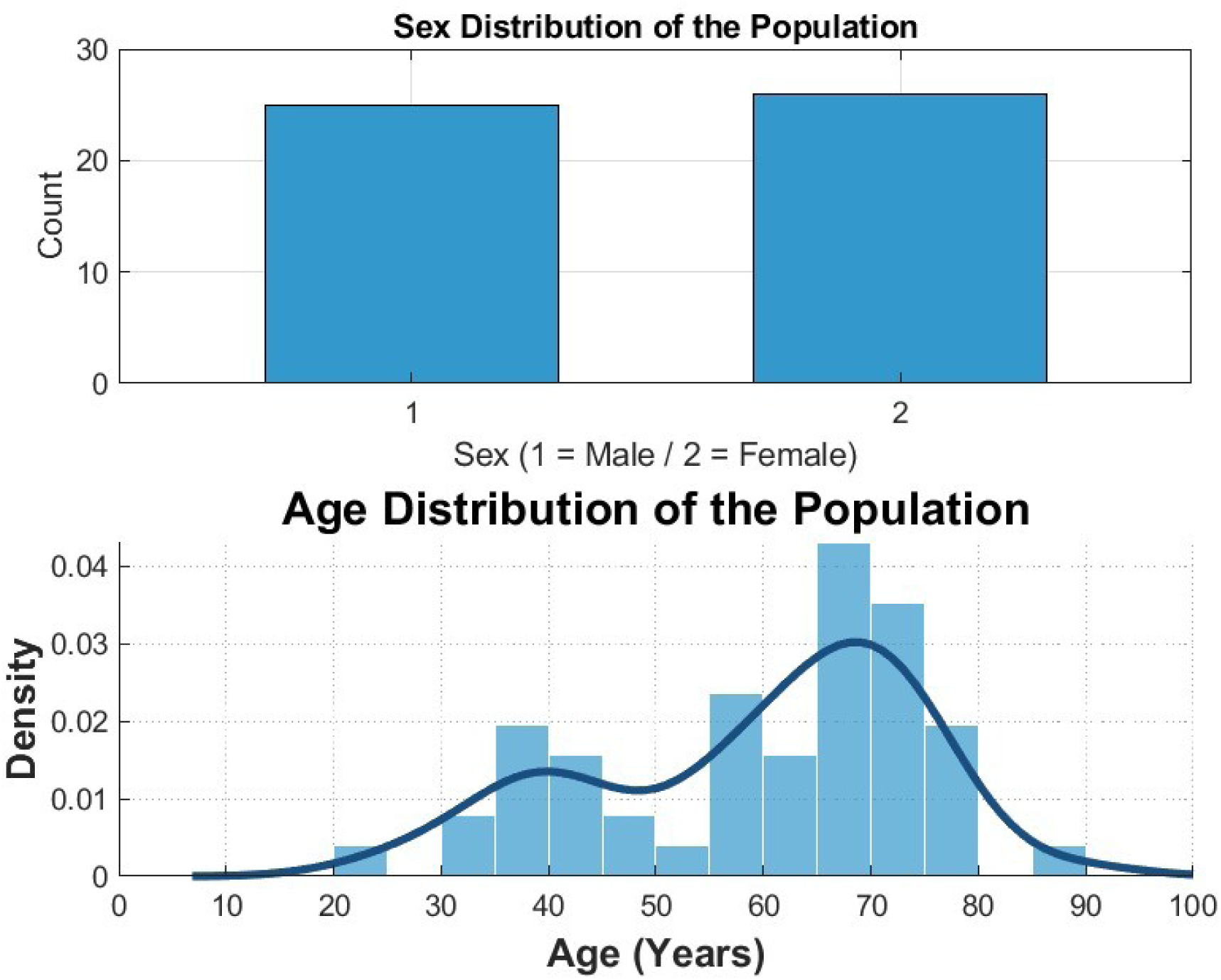
MGnet Telemedicine study

**Figure A4.**
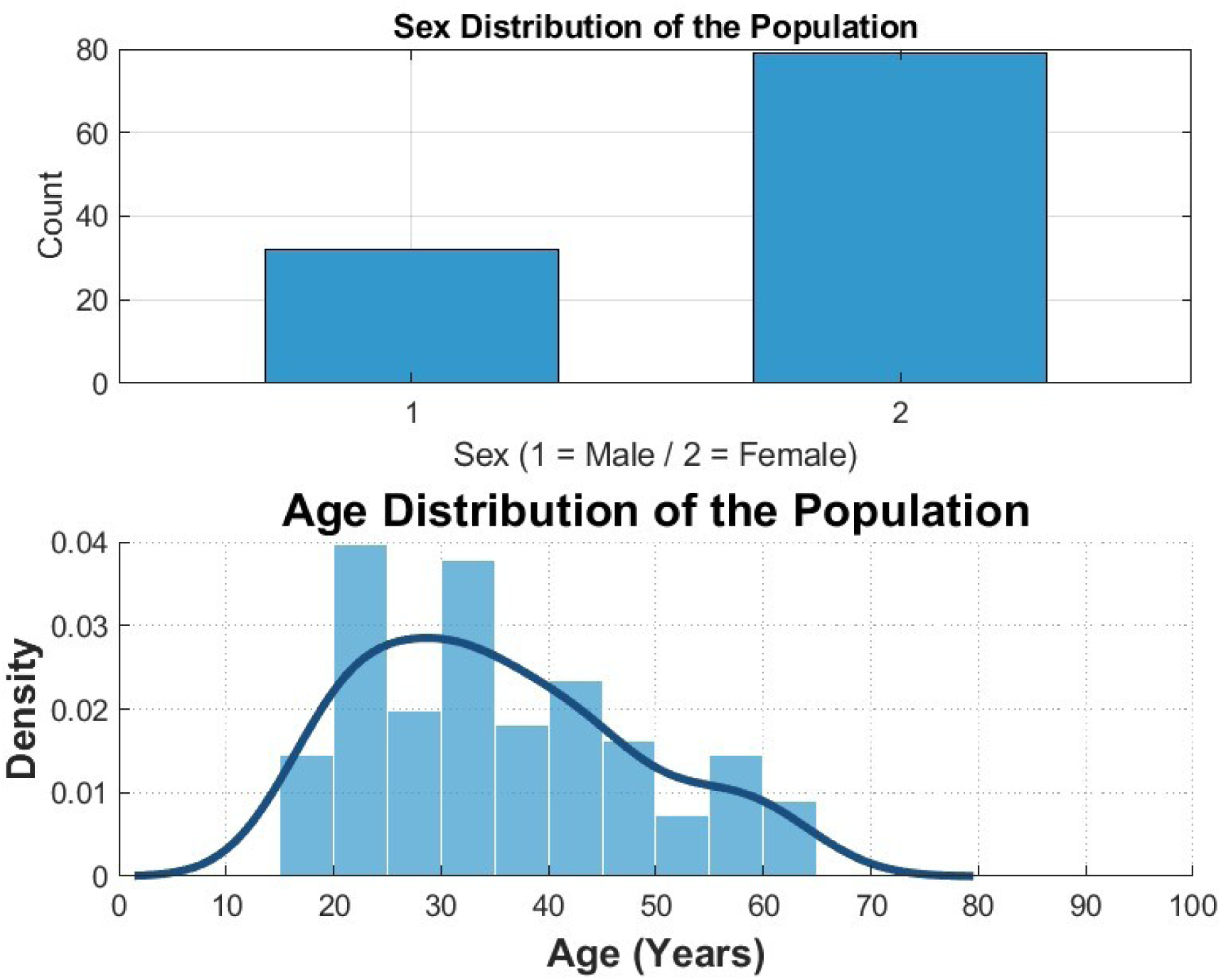
MGTX Clinical Trial

**Figure A5.**
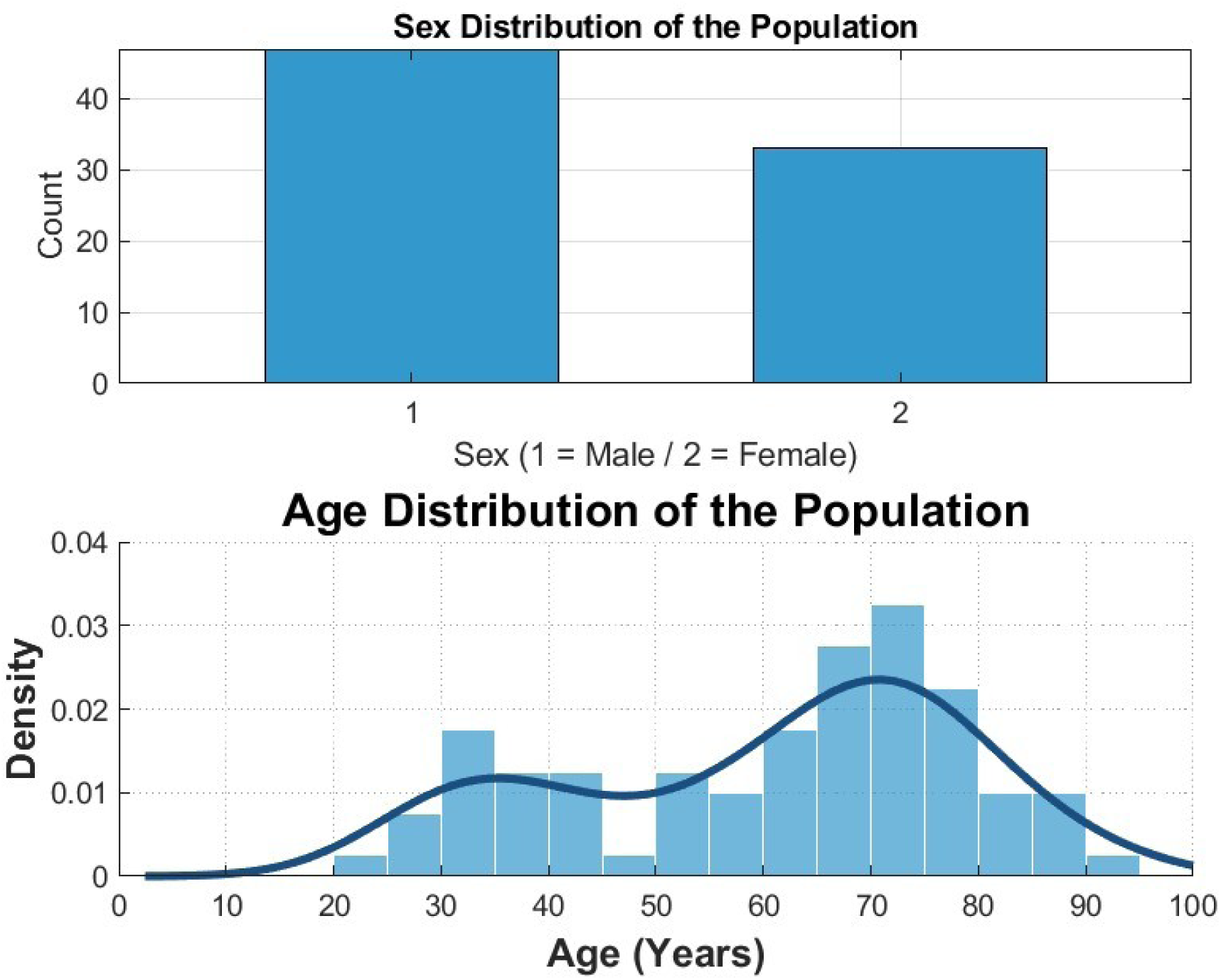
MSG Clinical Trial

**Figure A6.**
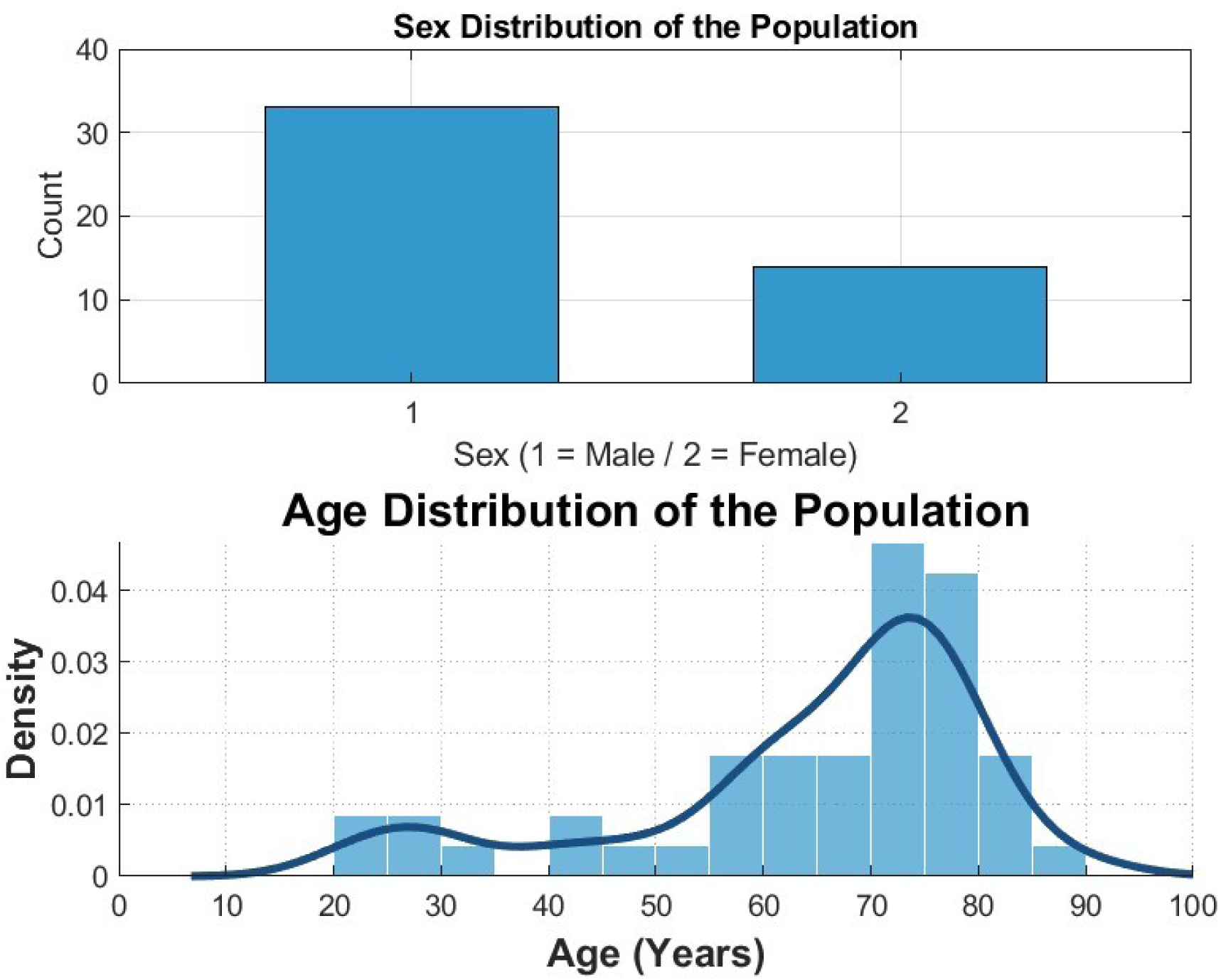
Rinomax Clinical Trial.

**Figure A7.**
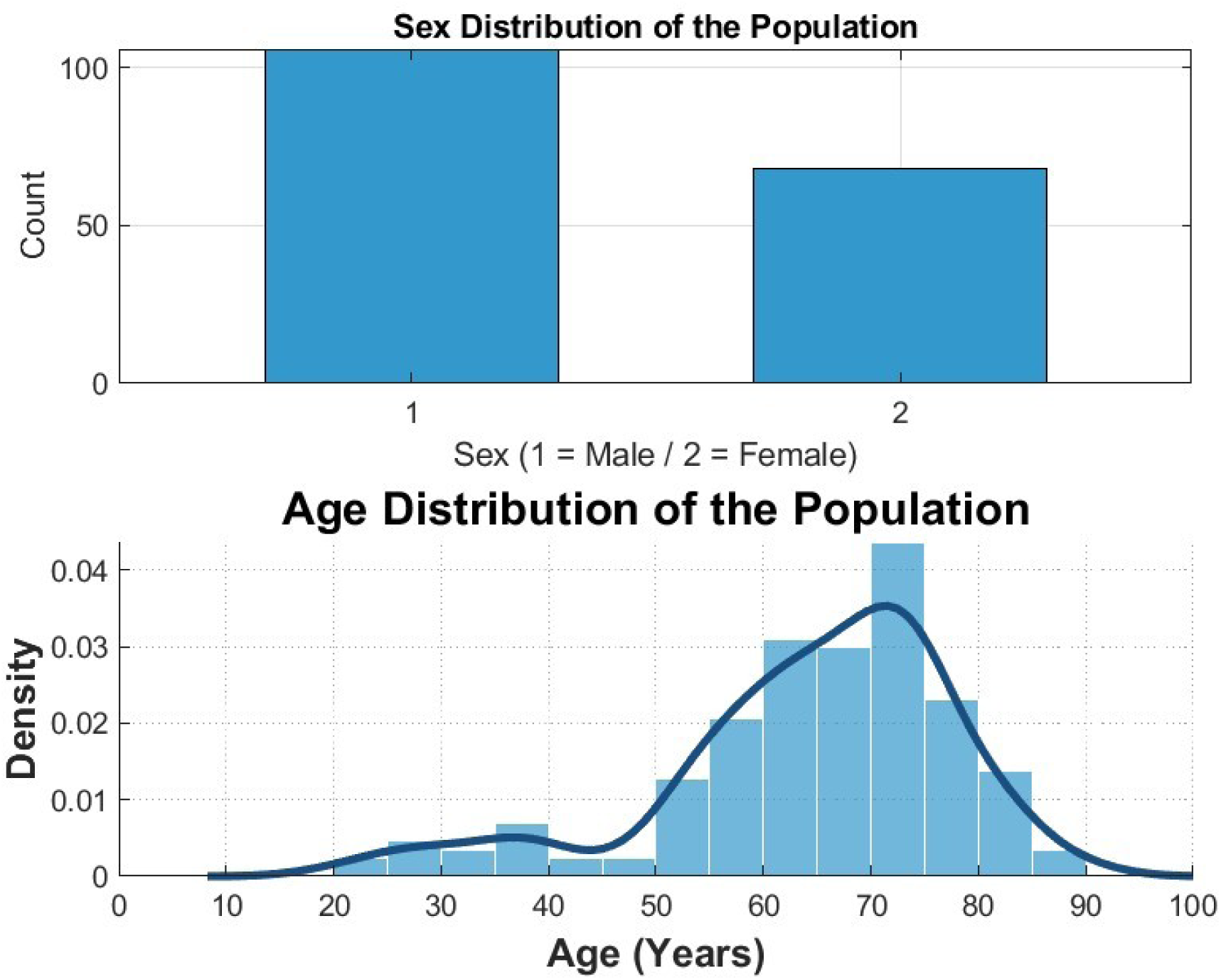
Promise Clinical Trial.

**Figure A8.**
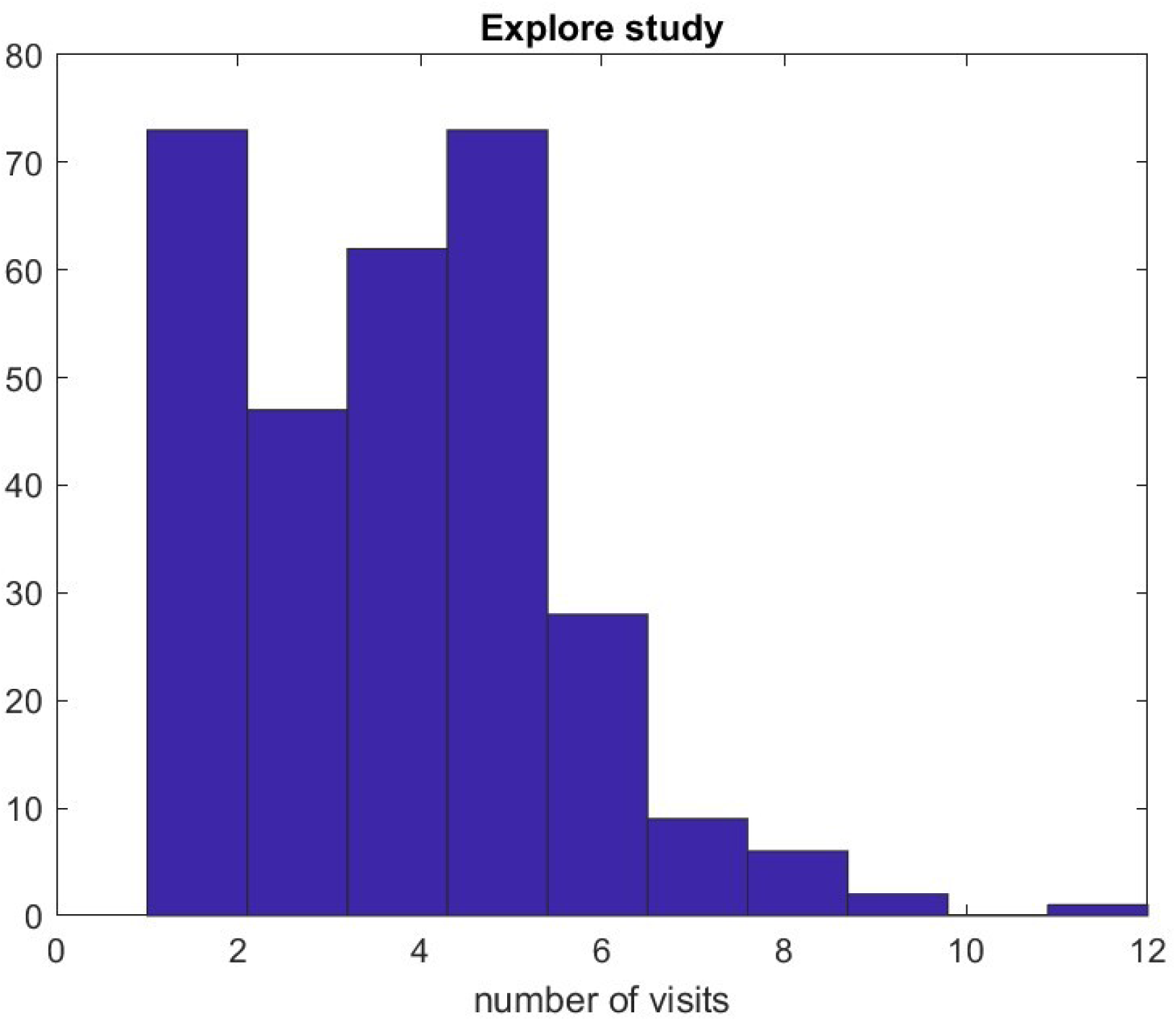
Distribution of the number of visits per patient in Explore.

**Figure A9.**
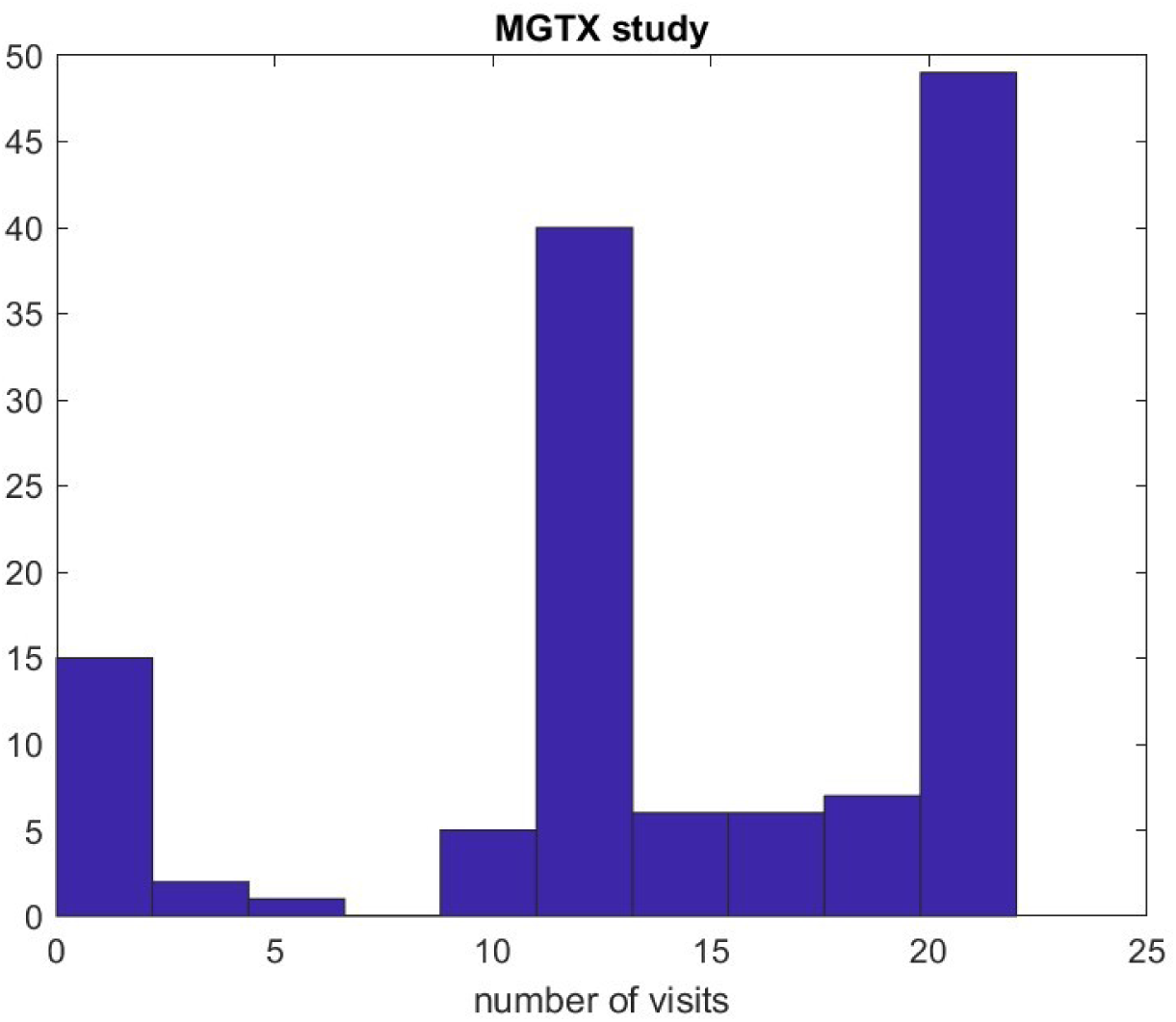
Distribution of the number of visits per patient in MGTX.

**Table A10.**
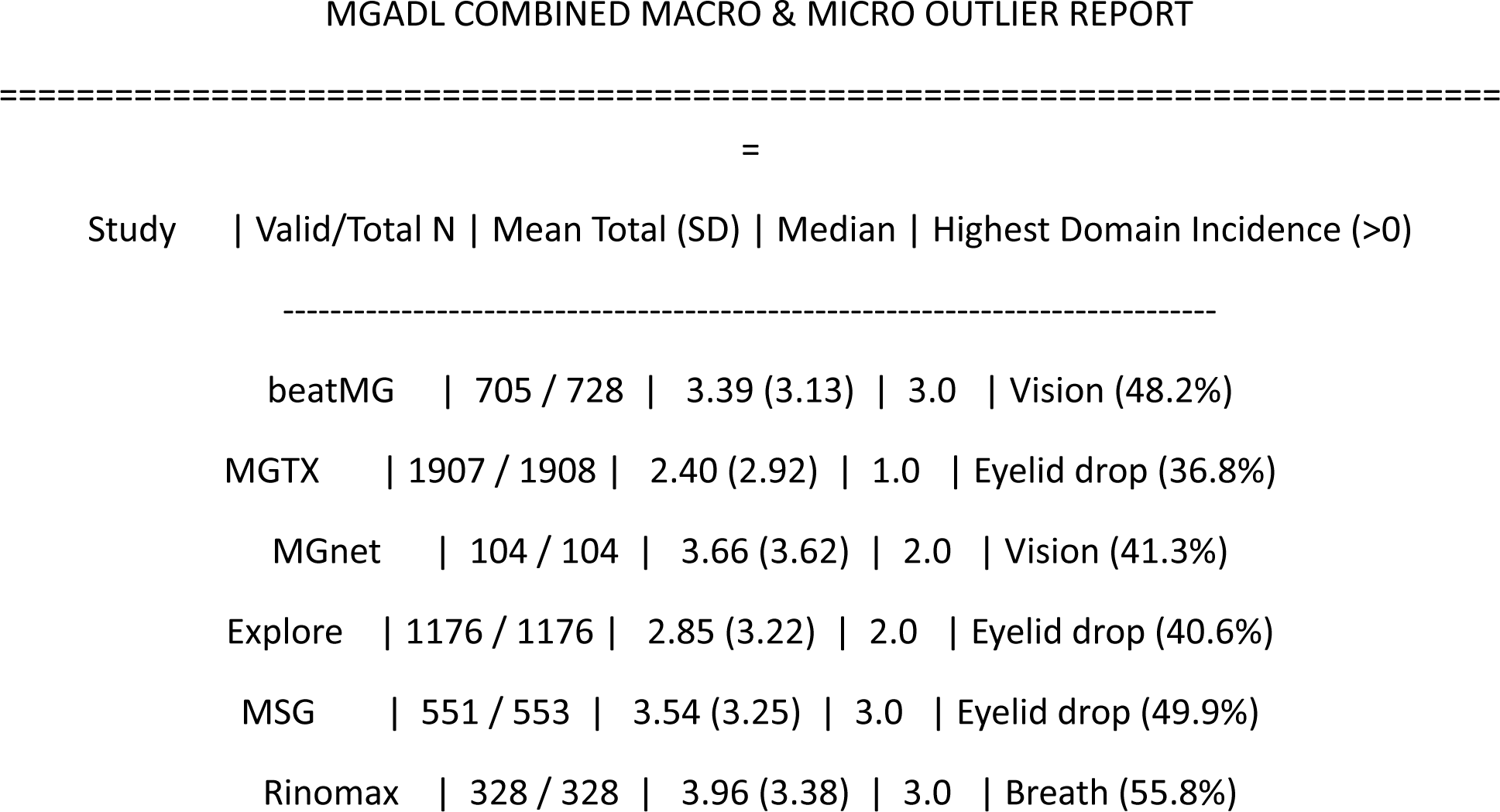
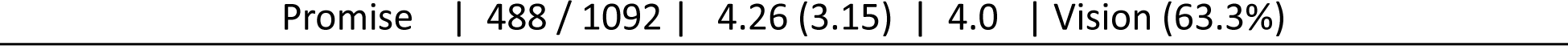

**Figure A11.**
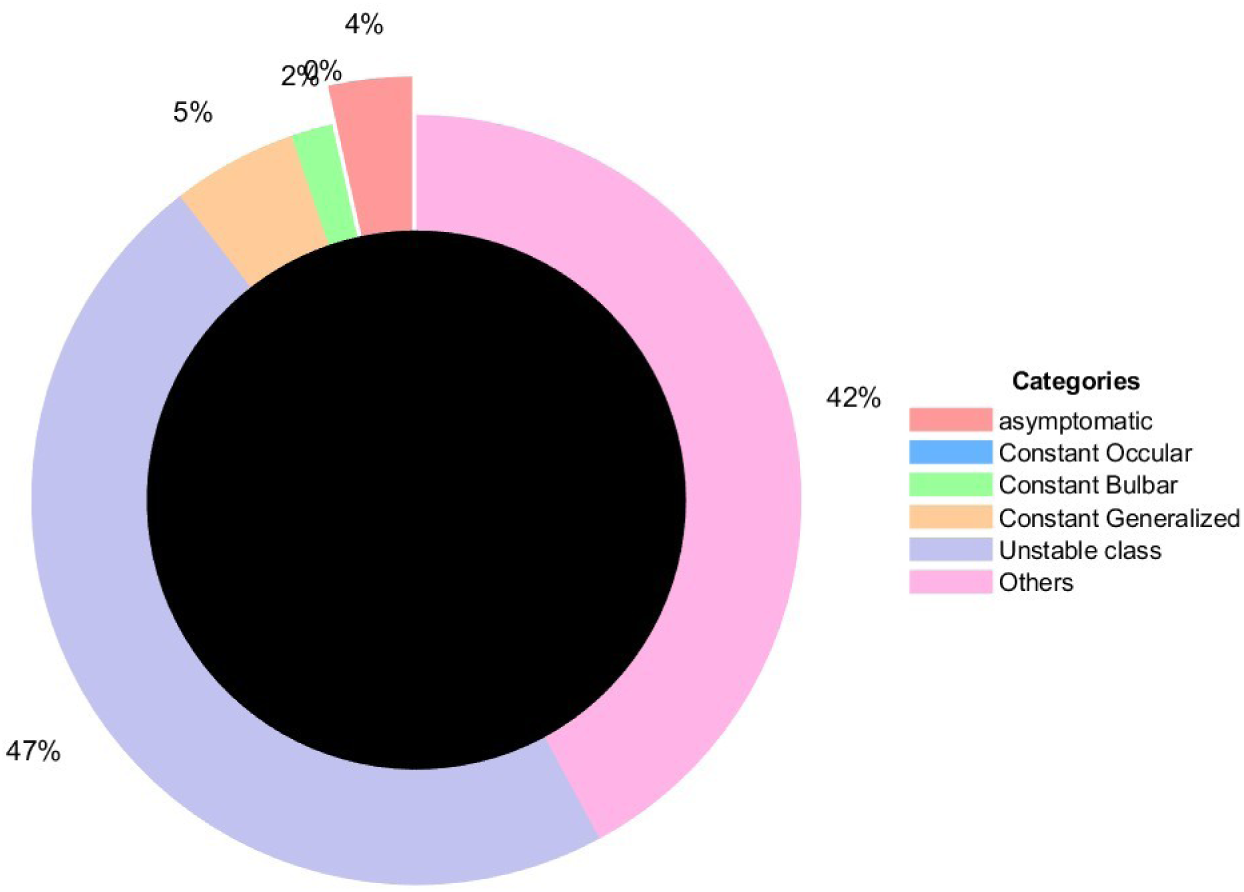
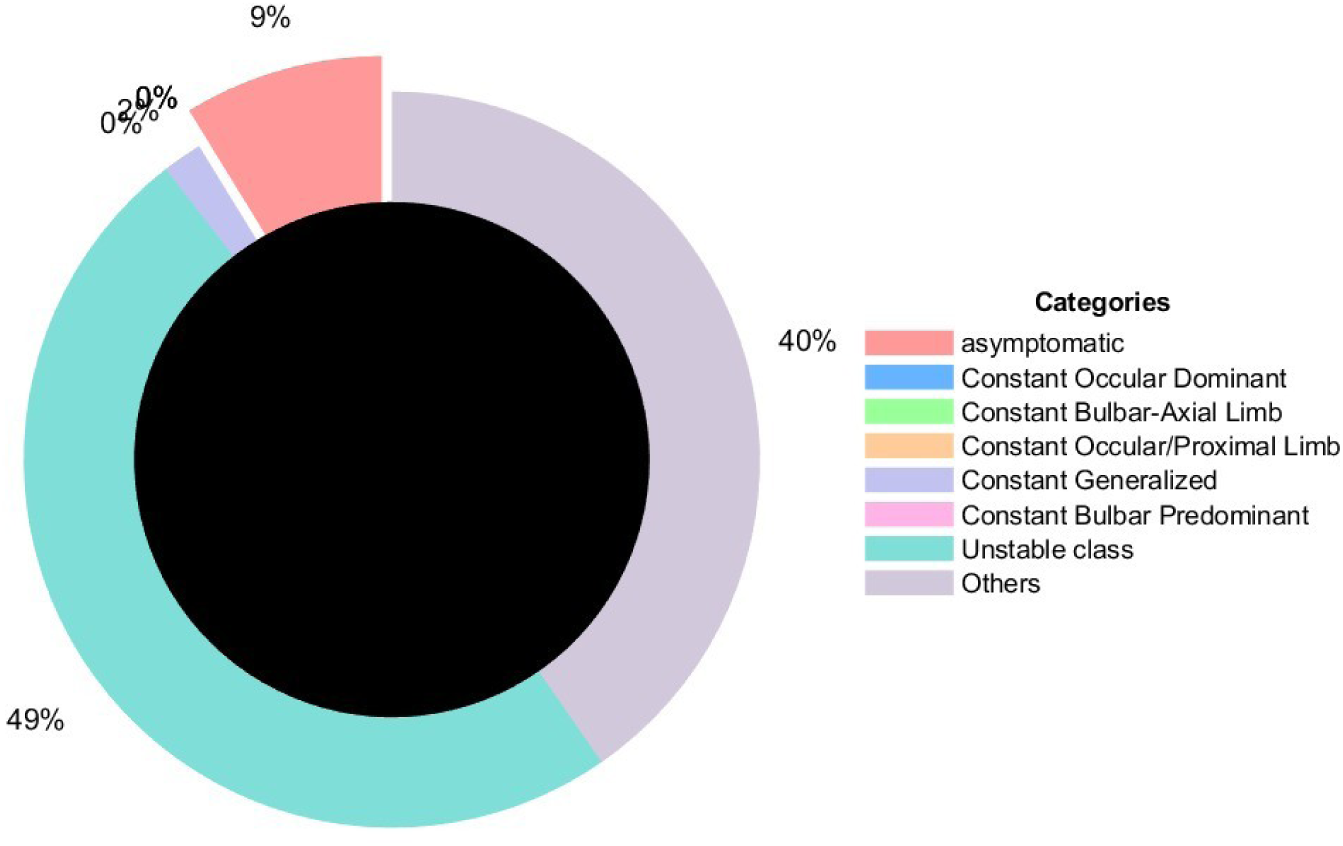
Stability of patient phenotypes in PROMISE, using the three-cluster model (top) and the five-cluster model (bottom). Approximately 47%–49% of patients had unstable longitudinal MG-ADL phenotypes, defined as a change in cluster assignment at least once over three visits. The analysis was restricted to patients with at least four visits, which represented the large majority of the cohort. Instability was more frequent than in MGTX and Explore.

**Figure A12.**
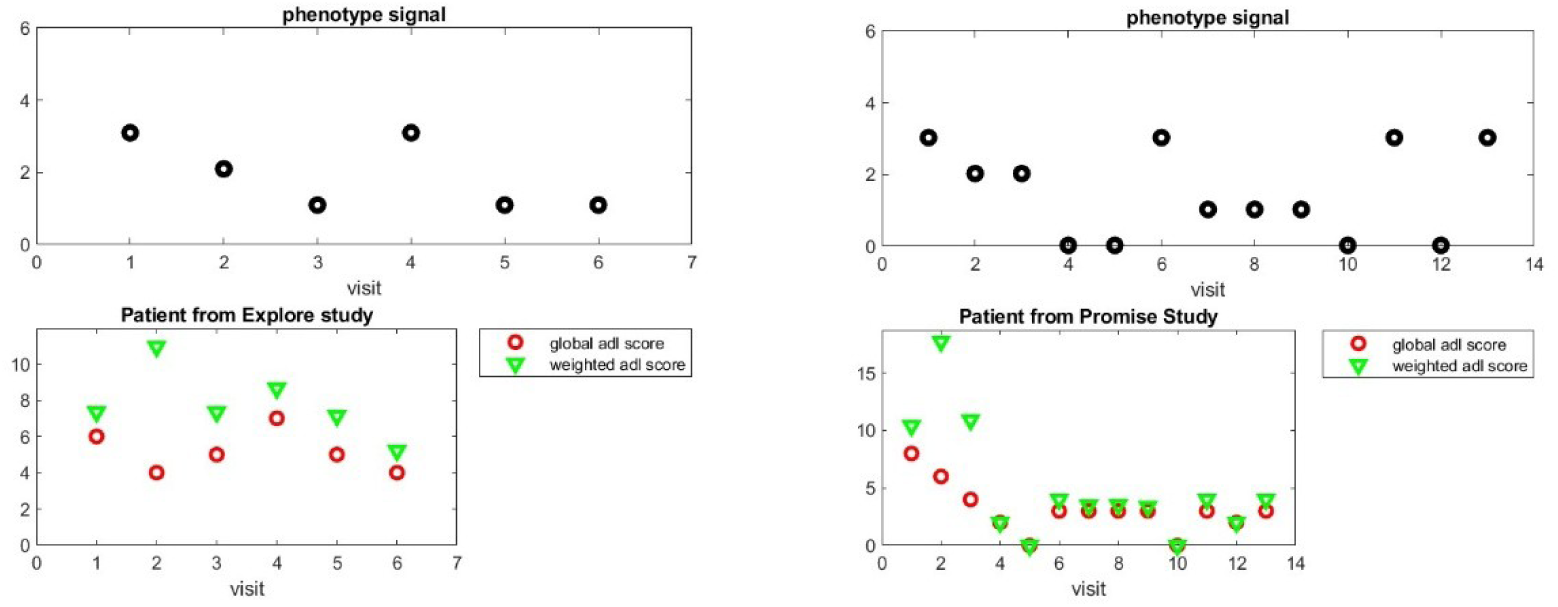
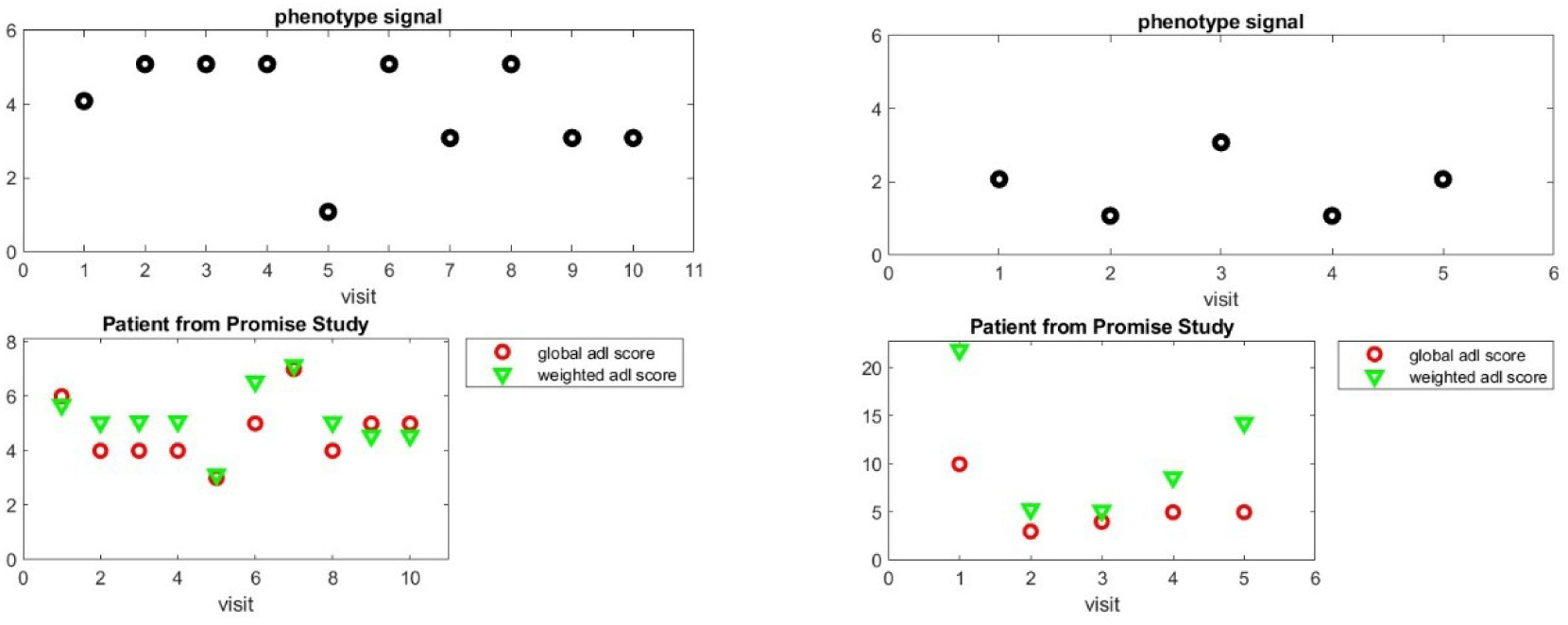
Selected trajectories illustrate repeated transitions between phenotypes. In each example, changes in the aggregate score over time can differ from changes in the weighted score. By design, the weighted score is more sensitive to redistribution among individual items, because small item-level changes may have a substantial effect even when the aggregate score changes little.

**Figure A13.**
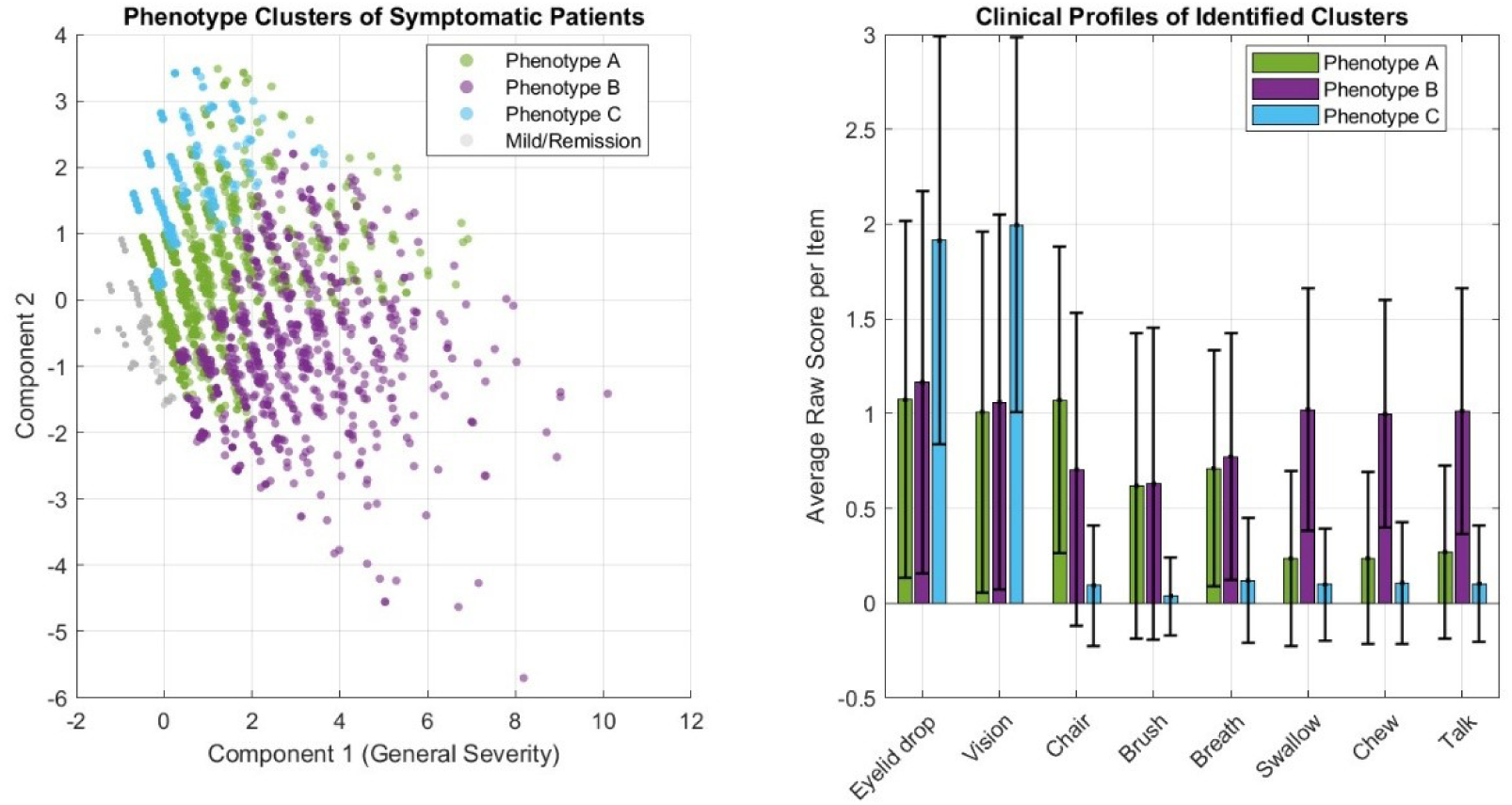
Classification using three clusters. The clusters corresponded broadly to ocular-dominant, bulbar-dominant, and generalized MG. This model did not separate the contributions of other items identified by PCA, including chair, brush, and breathing.

**Figure A14.**
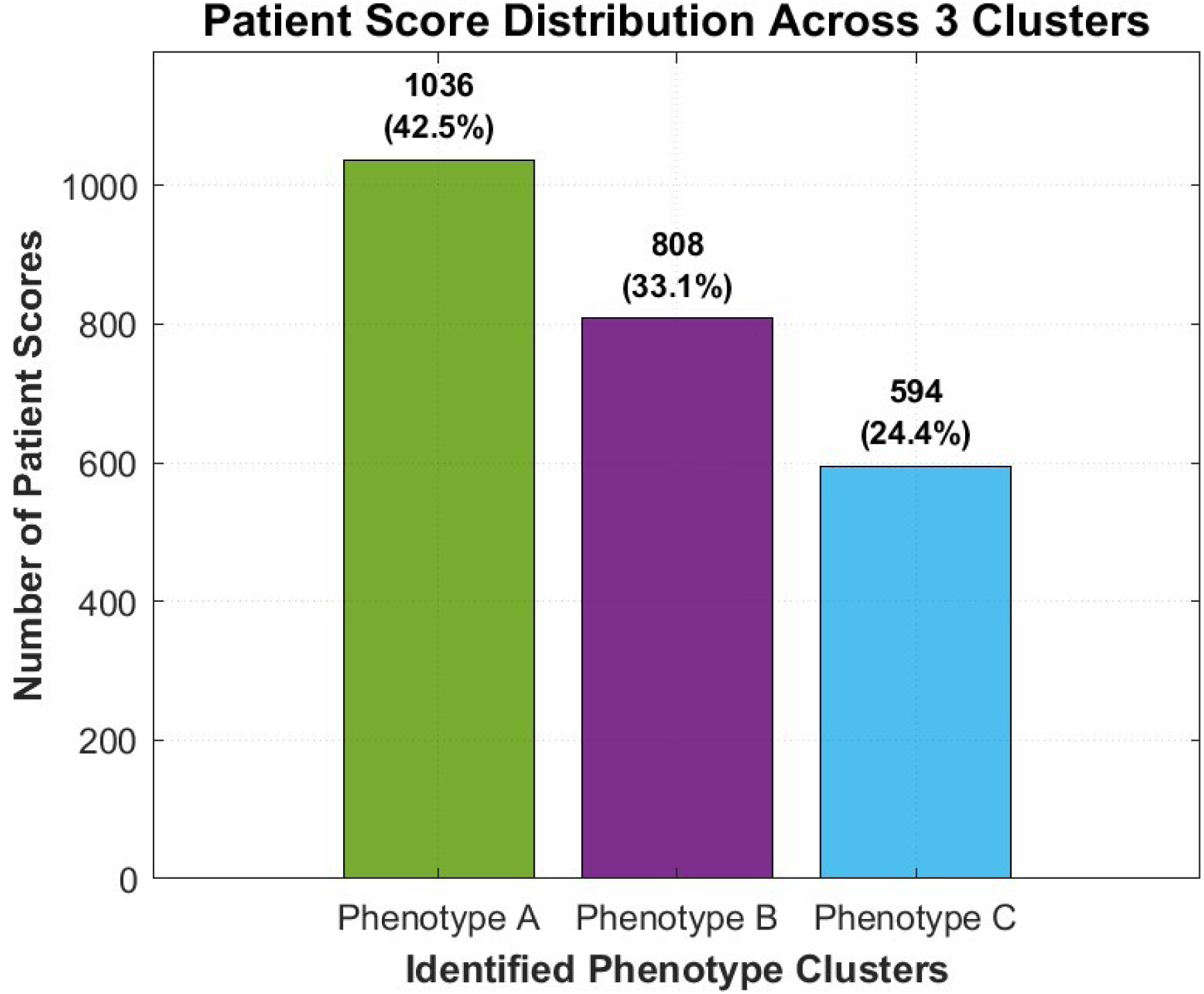
Distribution of symptomatic assessments across the three clusters.

## References

1. Stephenson D, Belfiore-Oshan R, Karten Y, Keavney J, Kwok DK, Martinez T, et al. Transforming Drug Development for Neurological Disorders: Proceedings from a Multidisease Area Workshop. Neurotherapeutics : the journal of the American Society for Experimental NeuroTherapeutics. 2023;20(6):1682–91.

2. Cedarbaum JM, Stephenson D, Rudick R, Carrillo MC, Stebbins G, Kerr D, et al. Commonalities and challenges in the development of clinical trial measures in neurology. Neurotherapeutics : the journal of the American Society for Experimental NeuroTherapeutics. 2015;12(1):151–69.

3. Jutten RJ, Papp KV, Hendrix S, Ellison N, Langbaum JB, Donohue MC, et al. Why a clinical trial is as good as its outcome measure: A framework for the selection and use of cognitive outcome measures for clinical trials of Alzheimer’s disease. Alzheimer’s & dementia : the journal of the Alzheimer’s Association. 2023;19(2):708–20.

4. Kaminski HJ, Sikorski P, Coronel SI, Kusner LL. Myasthenia gravis: the future is here. J Clin Invest. 2024;134(12).

5. Thomsen JLS, Andersen H. Outcome Measures in Clinical Trials of Patients With Myasthenia Gravis. Front Neurol. 2020;11:596382.

6. Wolfe GI, Herbelin L, Nations SP, Foster B, Bryan WW, Barohn RJ. Myasthenia gravis activities of daily living profile. Neurology. 1999;52(7):1487–9.

7. Janssen MF, Dewilde S, Wolfe GI, Muppidi S, Phillips G. Psychometric properties of MG-ADL items and MG-ADL score: An assessment of distributional characteristics, validity and factor structure in two large datasets. J Neurol Sci. 2024;463:123135.

8. McPherson T, Aban I, Duda PW, Farzaneh-Far R, Wolfe GI, Kaminski HJ, et al. Correlation of Quantitative Myasthenia Gravis and Myasthenia Gravis Activities of Daily Living scales in the MGTX study. Muscle Nerve. 2020;62(2):261–6.

9. Benatar M, Cutter G, Kaminski HJ. The best and worst of times in therapy development for myasthenia gravis. Muscle Nerve. 2023;67(1):12–6.

10. Group MS. A trial of mycophenolate mofetil with prednisone as initial immunotherapy in myasthenia gravis. Neurology. 2008;71(6):394–9.

11. Wolfe GI, Kaminski HJ, Aban IB, Minisman G, Kuo HC, Marx A, et al. Randomized Trial of Thymectomy in Myasthenia Gravis. N Engl J Med. 2016;375(6):511–22.

12. Nowak RJ, Coffey CS, Goldstein JM, Dimachkie MM, Benatar M, Kissel JT, et al. Phase 2 Trial of Rituximab in Acetylcholine Receptor Antibody-Positive Generalized Myasthenia Gravis: The BeatMG Study. Neurology. 2022;98(4):e376–e89.

13. Piehl F, Eriksson-Dufva A, Budzianowska A, Feresiadou A, Hansson W, Hietala MA, et al. Efficacy and Safety of Rituximab for New-Onset Generalized Myasthenia Gravis: The RINOMAX Randomized Clinical Trial. JAMA Neurol. 2022;79(11):1105–12.

14. Narayanaswami P, Sanders DB, Thomas L, Thibault D, Blevins J, Desai R, et al. Comparative effectiveness of azathioprine and mycophenolate mofetil for myasthenia gravis (PROMISE-MG): a prospective cohort study. Lancet Neurol. 2024;23(3):267–76.

15. Jolliffe IT, Cadima J. Principal component analysis: a review and recent developments. Philos Trans A Math Phys Eng Sci. 2016;374(2065):20150202.

16. Kaufman L, Rousseeuw P. Clustering by means of Medoids. In: Dodge Y, editor. Statistical Data Analysis Based on the L1-Norm and Related Methods: North-Holland; 1987. p. 405–16.

17. Cha SH. Comprehensive survey on distance/similarity measures between probability density functions. Int J Math Model Meth Appl Sci. 2007;1:300–7.

18. Meisel A, Sacca F, Spillane J, Vissing J, Sub-committee MGC. Expert consensus recommendations for improving and standardising the assessment of patients with generalised myasthenia gravis. Eur J Neurol. 2024;31(7):e16280.

19. Muppidi S, Silvestri NJ, Tan R, Riggs K, Leighton T, Phillips GA. Utilization of MG-ADL in myasthenia gravis clinical research and care. Muscle Nerve. 2022;65(6):630–9.

20. Guptill JT, Benatar M, Granit V, Habib AA, Howard JF, Jr., Barnett-Tapia C, et al. Addressing Outcome Measure Variability in Myasthenia Gravis Clinical Trials. Neurology. 2023;101(10):442–51.

21. Benatar M, Kaminski HJ, Quality Standards Subcommittee of the American Academy of N. Evidence report: the medical treatment of ocular myasthenia (an evidence-based review): report of the Quality Standards Subcommittee of the American Academy of Neurology. Neurology. 2007;68(24):2144–9.

22. Benatar M, Sanders DB, Burns TM, Cutter GR, Guptill JT, Baggi F, et al. Recommendations for myasthenia gravis clinical trials. Muscle Nerve. 2012;45(6):909–17.

23. Muppidi S, Wolfe GI, Conaway M, Burns TM, Mg C, Mg-Qol15 Study G. MG-ADL: still a relevant outcome measure. Muscle Nerve. 2011;44(5):727–31.

24. Jaretzki A, 3rd, Barohn RJ, Ernstoff RM, Kaminski HJ, Keesey JC, Penn AS, et al. Myasthenia gravis: recommendations for clinical research standards. Task Force of the Medical Scientific Advisory Board of the Myasthenia Gravis Foundation of America. Neurology. 2000;55(1):16–23.

25. Li HF, Gilhus NE, Yang H, Chen X. Editorial: Phenotypes of myasthenia gravis. Front Neurol. 2022;13:1025183.

26. de Meel RHP, Tannemaat MR, Verschuuren J. Heterogeneity and shifts in distribution of muscle weakness in myasthenia gravis. Neuromuscul Disord. 2019;29(9):664–70.

27. Khani-Habibabadi F, Verhamme FM, Moshir M, Casneuf T, Rao VTS, Steeland S, et al. Pathogenic properties of myasthenia gravis AChR autoantibodies associate with clinical response to efgartigimod. J Neurol Neurosurg Psychiatry. 2026;97(10):923–35.

28. Garbey M, Lesport Q, Kaminski HJ. Construction of patient trajectories to model clinical trial outcomes: application to myasthenia gravis. Front Digit Health. 2026;8:1755031.

29. Garbey M, Lesport Q, Kaminski HJ. Bridging the Heterogeneity of Myasthenia Gravis Scores as a Foundational Step Towards the Construction of a Digital Twin. Biomedicines. 2025;13(12).

